# A Multi-Symptom Circuit Architecture of Obsessive–Compulsive Disorder

**DOI:** 10.64898/2025.12.14.25342121

**Authors:** Barbara Hollunder, Garance M. Meyer, Ningfei Li, Pablo Reinhardt, Ilkem Aysu Sahin, Nanditha Rajamani, Julianna Pijar, Cristina Nombela, Philip Mosley, Nicola Acevedo, Kara A. Johnson, Nicole Provenza, Harith Akram, Benjamin M. Borron, Tina Chou, Jürgen Germann, Juan Pablo Castaño Montoya, Bryan Strange, Juan A. Barcia, Himanshu Tyagi, David J. Castle, Susan Rossell, Peter Bosanac, Carsten Finke, Andrea A. Kühn, Domenico Servello, Alberto R. Bona, Mauro Porta, Alon Y. Mogilner, Michael H. Pourfar, Jill L. Ostrem, Jens Kuhn, Veerle Visser-Vandewalle, Stephan Chabardes, Sameer A. Sheth, Wayne K. Goodman, Andrew H. Smith, Ki Sueng Choi, Brian H. Kopell, Helen S. Mayberg, Martijn Figee, Thomas Foltynie, Patricia Limousin, Linda Ackermans, Joost J. A. de Jong, Albert F. G. Leentjens, Christopher R. Butson, Michael S. Okun, R. Mark Richardson, G. Rees Cosgrove, Darin D. Dougherty, Shan H. Siddiqi, Andres M. Lozano, Ludvic Zrinzo, Eileen Joyce, Mircea Polosan, Clemens Neudorfer, Juan Carlos Baldermann, Michael D. Fox, Andreas Horn

## Abstract

Obsessive–compulsive disorder (OCD) manifests with diverse symptom constellations that likely arise from dysfunction in partially distinct neural circuits. Deep brain stimulation (DBS), paired with high-resolution connectomics, offers a unique window into these pathways in humans. Here, we analyzed clinical outcomes and stimulation sites from 77 treatment-refractory patients with OCD and 39 with Tourette’s syndrome (TS) exhibiting comorbid obsessive–compulsive behaviors (244 electrodes across 15 cohorts and eight targets). Engagement of the previously defined *OCD response tract* predicted global obsessive-compulsive symptom improvement across heterogeneous targets and diagnoses, spanning both OCD and TS. Beyond broad symptoms, obsessions, compulsions, anxiety, and depression mapped onto distinct yet overlapping subcircuits fragmenting the anterior limb of the internal capsule along a dorsoventral axis. This fine-scale architecture of OCD circuit dysfunction was reproducible across cross-validation schemes and patient subsets. Exploratory analyses identified additional subcircuits for cognitive control and flexibility. Global functional recovery was better explained by combined engagement of multiple symptom-specific rather than a single tract. Collectively, these findings illustrate how invasive neuromodulation can delineate a multi-symptom circuit taxonomy of compulsivity that may guide personalized neuromodulation.

## Introduction

A key challenge in treating obsessive-compulsive disorder (OCD), a common psychiatric condition with a lifetime prevalence of 2-3%^1^, lies in its multifaceted symptom profile^2^. Patients may experience diverse and unique combinations of intrusive, distressing thoughts (obsessions) paired with ritualistic behaviors or mental acts (compulsions) that can consume significant time and energy^1,3,4^. In addition, OCD frequently presents with secondary symptoms or comorbidities^1,2,4–7^, including depression and anxiety, which collectively impair daily functioning and socio-occupational engagement^8^. Obsessive-compulsive behaviors also commonly arise in tic disorders such as Tourette’s syndrome (TS)^9^, and this overlap may reflect a shared compulsivity spectrum^4,5,10,11^.

Decades of neuroimaging work have sought to anchor this heterogeneity in circuit-level dysfunction^2,4–6,12^. Early circuit models inspired by these findings centered on imbalances within cortico–striato–thalamo–cortical loops, proposing that hyperactive ventral affective and hypoactive dorsal cognitive pathways drive the combination of anxiety, cognitive rigidity, and repetitive behavior^13^. More recent frameworks have expanded to a multi-circuit architecture proposing fronto-limbic (fear regulation), ventral affective (reward processing), dorsal (executive control and emotion regulation) and ventral cognitive (self-regulation and motor inhibition), as well as sensorimotor circuit components (motor control)^2,5^.

Therapeutic limitations underline the urgency of bridging the gap between such conceptual models and empirical evidence: ∼10% of individuals with OCD fail to respond to first-line interventions including cognitive-behavioral or pharmacotherapy^14,15^, prompting the advent of deep brain stimulation (DBS)^16^. Yet after two decades and numerous putative targets, the field still continues to debate the “optimal” site of stimulation^17^. Early efforts were inspired by robust symptom relief after ablative procedures, comprising capsulotomy of the anterior limb of the internal capsule (ALIC) and cingulotomy of the anterior cingulate cortex (ACC)^18,19^. Consequently, several ventral capsule/ventral striatum (VC/VS) targets were probed, including the ventral ALIC^16,20,21^ or the VC/VS^22^ with the nucleus accumbens (NAcc)^23,24^. Other suggested targets comprised the bed nucleus of the stria terminalis (BNST)^25–28^, forming part of the extended amygdala, and the inferior thalamic peduncle (ITP)^29^, the ventral-most extension of the VC/VS circuitry connecting the thalamus and orbitofrontal cortex (OFC). Serendipitous observations of alleviated OCD and cognitive-affective (side-)effects in Parkinson’s disease later inspired several targets within the subthalamic region^30^: the anteromedial subthalamic nucleus (STN)^31^ and the ventral tegmental area projection pathway (also denominated superolateral medial forebrain bundle, sl-MFB)^32^.

Intriguingly, similar OCD response for ∼60% of patients across these targets^17,33,34^ implied that the beneficial effects of DBS may hinge less on a singular anatomical location and more on engagement of a shared circuit^35–39^. Indeed, connectomic analyses identified a fronto-subcortical tract associated with optimal therapeutic DBS response of global obsessive-compulsive symptoms, as measured by the Yale-Brown Obsessive-Compulsive Scale (Y-BOCS)^36^. Receiving projections from various prefronto-cortical regions, this *OCD response tract* interconnects a broad set of anatomical targets as it courses through the central ALIC and sends projections likely to STN and other midbrain and brainstem structures^38,40^. Growing evidence links stimulation of this normative response tract with optimal group-level DBS response, as independently replicated across multiple surgical targets — including ALIC^41–43^, BNST^26^, internal pallidum^44^, and sl-MFB regions^45^ (for reviews, see^46,47^). A blinded validation study^43^, as well as successful targeting and DBS reprogramming in individual cases^39,48,49^ have further cemented the OCD response tract as a promising therapeutic axis.

Importantly, however, this OCD response tract has been defined (i) at the group level, and (ii) tied to the full symptom composite captured by the Y-BOCS. Hence, this approach collapses individual symptom dimensions (obsessions vs. compulsions) and overlooks common secondary symptoms or comorbid features such as anxiety and depression^50^. At the same time, the internal capsule itself exhibits a highly ordered topography with ventrodorsal and mediolateral gradients reflecting distinct prefrontal inputs^38^. This anatomical organization may carry important functional implications for OCD treatment: distinct circuit elements, each associated with specific symptom domains, could be preferentially recruited as a function of the precise DBS electrode placement within the ALIC. However, this relationship has yet to be empirically tested. Linking the ALIC’s fiber architecture to nuanced symptom outcomes could both help further curate OCD circuit models^2,5,13^ and guide personalized targeting^2,47,50,51^.

To address this gap, the present study leveraged a large multi-center, multi-target dataset of invasive DBS treatments for OCD (N = 77) and comorbid obsessive-compulsive behavior in TS (N = 39). We first tested whether engagement of the canonical fronto–subcortical OCD response tract^36^ would account for global obsessive–compulsive symptom relief irrespective of primary diagnosis and stereotactic target. We then investigated the relationship between select OCD symptoms (obsessions, compulsions, depression, anxiety) or cognitive features (dysfunction in cognitive flexibility and control) and associated normative tracts implicated in their DBS response. Finally, we integrated these findings into a multi-symptom circuit model to predict improvements in global functioning. Together, these results may shed light onto the heterogeneous circuit dysfunctions underlying compulsivity and chart a roadmap toward personalized brain-circuit therapeutics.

## Results

### Patient Demographics and Clinical Characteristics

*OCD Cohort:* The final cohort used to set up tract models included 77 patients with refractory OCD (40 females) from eight surgical centers who underwent bilateral DBS to one of six different stereotactic targets (ALIC, NAcc, STN, ITP, BNST, or VC/VS and STN combined). Among these patients, the average age at time of surgery was 38.2 ± 14.2 years, and they presented with an average age at the onset of OCD symptoms of 17.8 ± 10.2 years. Comprehensive demographic and clinical characteristics of patients are available in **Table S1** and in the original publications^26,29,35,52–55^. Across the patient sample, there was a notable effect of intervention on OCD symptoms, as assessed via the Y-BOCS, with an average improvement of 41.6% ± 24%. Notably, 54.6% of patients (n = 42) were classified as responders, defined by a Y-BOCS improvement of over 35% as suggested in the literature^33,34,56,57^. On a more nuanced level, average improvements in obsessions amounted to 39.7% ± 27.8% and those in compulsions to 41.2% ± 27.7%. In line with this, the impact of the intervention on patients’ everyday functioning was substantial, with a mean benefit of 110.4% ± 198.2% on the Global Assessment of Functioning (GAF). An overall improvement in comorbid symptoms was present, yet less pronounced, and characterized by high within-cohort variability: on average, patients improved by 25.3% ± 51.3% in depression, and by 17.9% ± 33.6% in anxiety.

As expected, most symptom improvements were also significantly correlated with one another. Specifically, there was a positive correlation between improvements in obsessions vs. compulsions (r = 0.59, p < 0.001), compulsions vs. anxiety (r = 0.43, p = 0.014), obsessions vs. anxiety (r = 0.4, p = 0.025), depression vs. anxiety (r = 0.37, p = 0.035), obsessions vs. depression (r = 0.33, p = 0.013), as well as compulsions vs. depression (r = 0.32, p = 0.015). These correlations, as discussed below, may both explain the overlaps across tract mappings among the four symptom clusters and substantiate the spatial segregation observed between them.

Further, only anxiety showed a significant association with global functioning (r = 0.44, p = 0.013), unlike its non-significant relationship to improvements in obsessions (r = 0.26, p = 0.098), compulsions (r = 0.2, p = 0.204), and depression (r = 0.21, p = 0.209).

*TS Cohort:* In addition to data of patients diagnosed with OCD, our analysis aimed at testing the validity of the published global OCD response tract^36^ as a target for treatment of obsessive-compulsive behavior irrespective of disease category and target. For this purpose, we included data from a seven-center cohort of N = 39 patients with TS (13 females) and comorbid obsessive-compulsive behavior, implanted with bilateral DBS electrodes either to the anteromedial internal pallidum (amGPi) (n = 24) or the centromedial thalamus (cmThal) (n = 15) from the International Tourette Syndrome DBS Database and Registry^58^ (https://tourettedeepbrainstimulationregistry.ese.ufhealth.org/). On average, TS patients improved by 18.7 % ± 49.0% on the Y-BOCS following DBS. **Table S2** comprises detailed information on demographic and clinical characteristics of the TS cohort.

**Fig. 1** summarizes applied methodological concepts. Electrode locations of the OCD cohort are visualized both for the full cohort as well as grouped by target in **Fig. 2**.

**Fig. 1.**
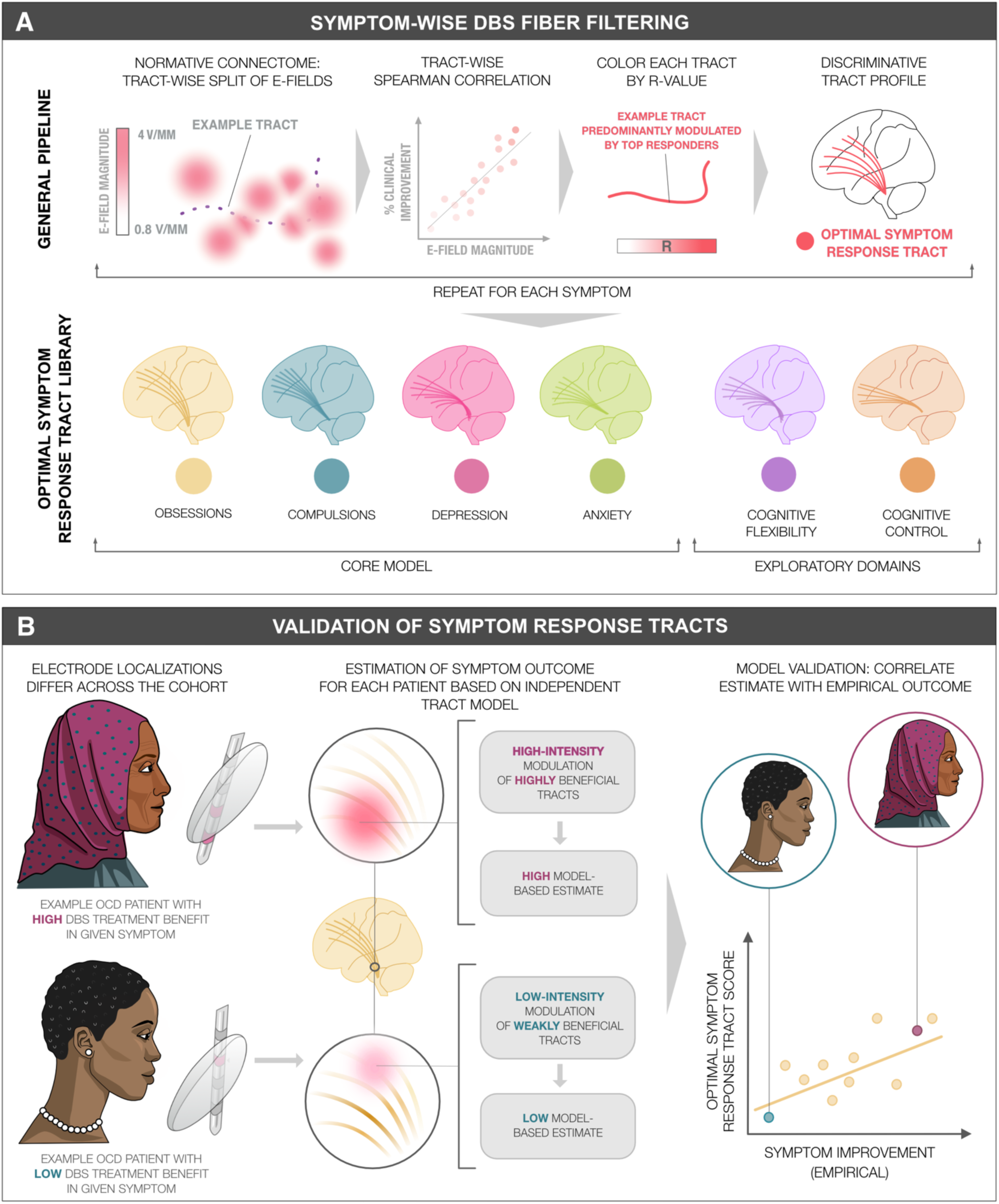
Methodological workflow for symptom response circuit modeling and validation. **(A)** Imaging-derived electrode placements were used to estimate electrical fields (E-fields) based on patient-specific stimulation parameters. Each tracts represented in a normative connectome was then assessed for symptom response when stimulated using established methods^39^. We first extracted the peak E-field magnitude sampled along each tract and correlated these values with percent symptom improvements for all intersecting E-fields using Spearman’s correlation. The resulting correlation coefficient was then used to weight and color-code each tract, with more intense colors indicating greater relevance for symptom improvement. Repeated across the connectome, this approach yielded an optimal connectivity model of tracts with therapeutic potential when stimulated. Applying this procedure across various symptoms, we established a library of response tracts linked to the most relevant dimensions in OCD. **(B)** We subsequently applied established methods to validate each symptom response tract for its capacity to estimate clinical outcomes, both within the cohort and in hold-out data^39^. To achieve this, we derived an outcome estimate based on E-field overlaps with the optimal symptom response tract model for each patient. Following previous reports^39^, we hypothesized that greater overlap would correlate with higher treatment benefits, while lower overlap would indicate comparably diminished outcomes. To validate this idea, we correlated these estimates with empirical symptom outcomes across the patient cohort.

**Fig. 2.**
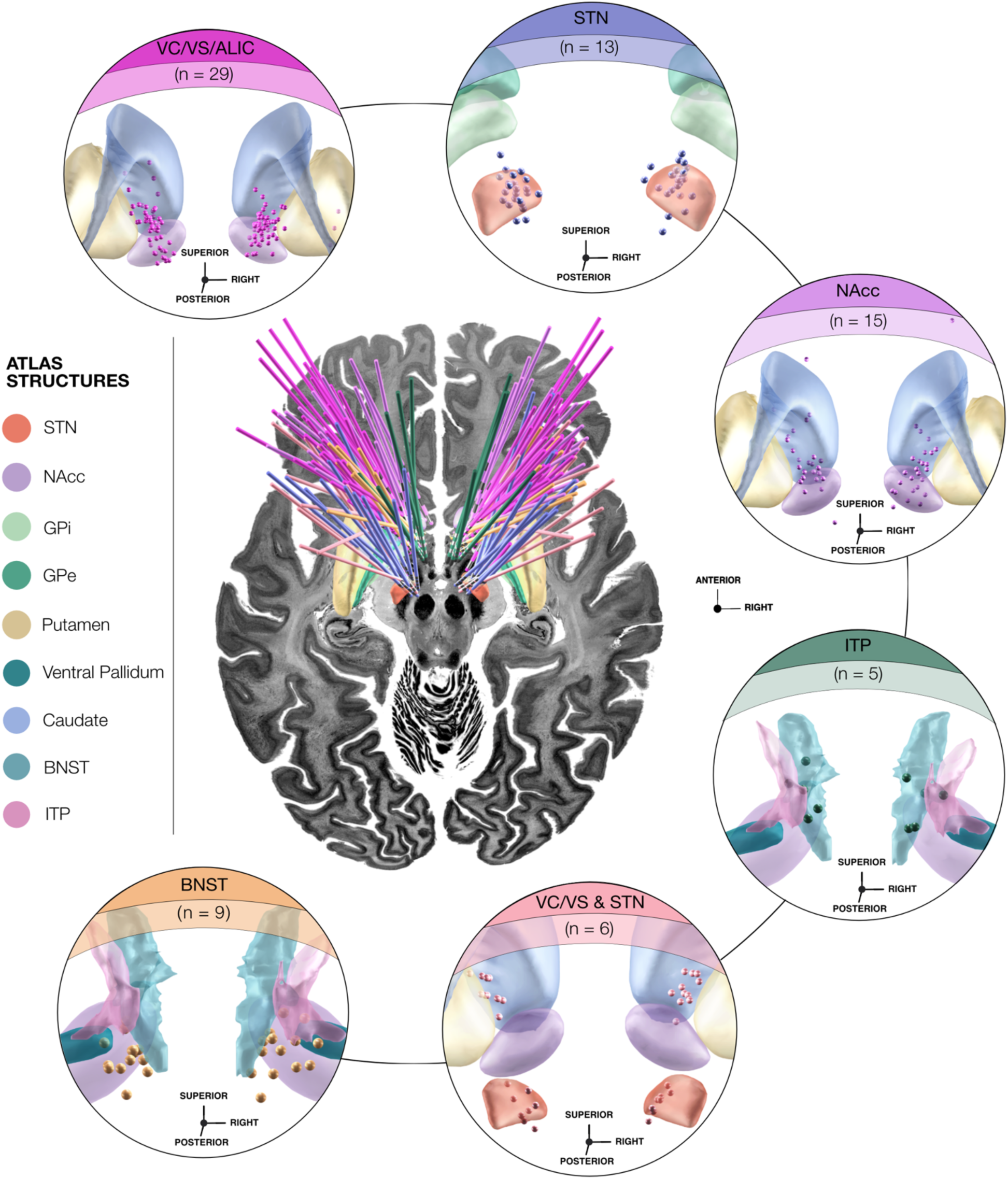
Diversity of electrode placements across the obsessive-compulsive disorder (OCD) patient cohort. *Center:* Bird-eye view of bilateral deep brain stimulation (DBS) electrode placements along with lead trajectories pooled across cohorts in anatomical context, superimposed on top of an axial plane (z = -10 mm) of the BigBrain template in 100 µm resolution^59^. *In-set circles:* Target-wise grouping of active electrode contacts, each in relation to adjacent neuroanatomical structures. Note that labeling reflects the intended target as reported in the respective original publication. The selected active contact may overlap with adjacent structures due to close spatial proximity. Subthalamic nucleus (STN), globus pallidus internus (GPi) and externus (GPe) as defined by the DBS Intrinsic Template (DISTAL) atlas^60^, version 1.1; the nucleus accumbens (NAcc), putamen, ventral pallidum, and caudate by the California Institute of Technology reinforcement learning atlas (CIT168) atlas^61^, version 1.1; and the bed nucleus of the stria terminalis (BNST) as well as the inferior thalamic peduncle (ITP) by the Atlas of the Human Hypothalamus^62^, version 1.0. *Abbreviations:* ALIC, anterior limb of the internal capsule; VC/VS, ventral capsule/ventral striatum.

### From Disease- to Symptom-Centricity: A Unified Response Circuit for Obsessive-Compulsive Symptoms

As a first step, we sought to validate the previously published OCD response tract^36^ in the present and larger clinical cohort. In contrast to the original tract definition^35,36^, which relied on a smaller and more anatomically homogeneous sample, our dataset encompassed 77 patients with considerable variability in DBS electrode placements. This variability stemmed from the inclusion of multiple international centers, stereotactic targets, and surgical differences in targeting strategies (**Fig. 2**). Even so, the degree of overlap between each patient’s stimulation volume and the previously identified OCD response tract remained a significant predictor of therapeutic efficacy. Specifically, greater tract modulation was associated with more pronounced improvements in global obsessive-compulsive symptoms captured by the Y-BOCS (R = 0.30, p = 0.009) (**Fig. 3**).

**Fig. 3.**
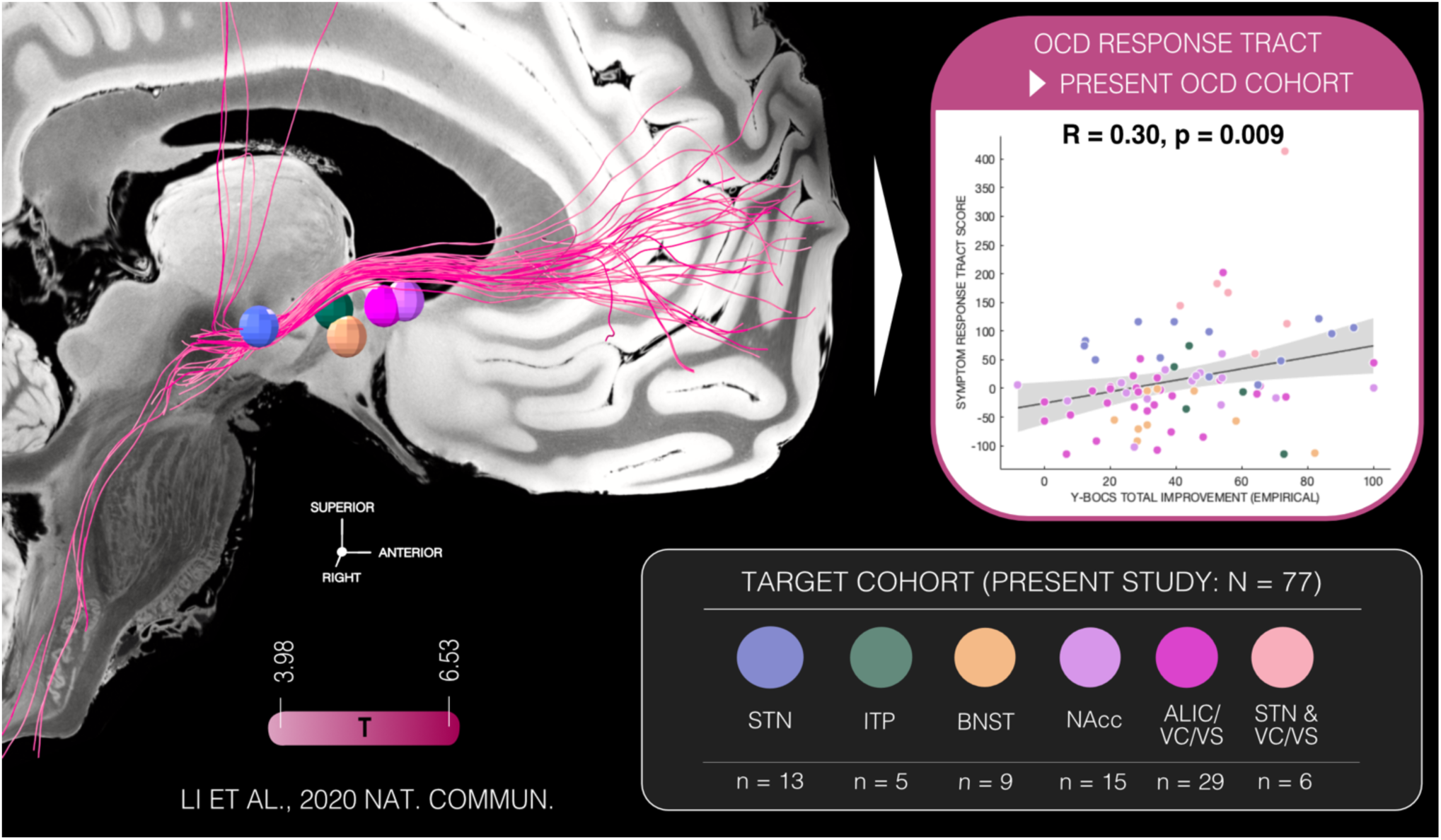
A unified deep brain stimulation (DBS) response tract for obsessive-compulsive symptoms. *Left.* The previously identified tract associated with optimal deep brain stimulation (DBS) response in global obsessive-compulsive disorder (OCD) symptomatology^36^, anchored to the Yale-Brown Obsessive-Compulsive Scale (Y-BOCS) Total score, is overlayed on top of a sagittal slice (x = -5 mm) from the 7T ex-vivo human brain atlas^63^. Individual tracts are color-coded according to T-value as described in Li et al.^36^, reflecting each tract’s ability to differentiate between good and poor DBS response. Average locations of activated electrode contacts from the N = 77 OCD patients are shown as spheres, color-coded by target group. Notably, London patients (n = 6) received stimulation at two distinct targets — the subthalamic nucleus (STN) and ventral capsule/ventral striatum (VC/VS). For visualization on the left, electrodes from these patients were categorized separately under each corresponding target. *Right.* The Spearman correlation plot demonstrates that stimulation volume overlap (represented as Symptom Response Tract Score attributed to each stimulation volume, averaged across bilateral estimates per patient) with the global OCD response tract defined by Li et al. (2020)^36^ significantly estimated patient responses in the current cohort (one-sided tests). The gray-shaded area represents the 95% confidence interval. *Abbreviations:* BNST, bed nucleus of the stria terminalis; ITP, inferior thalamic peduncle; NAcc, nucleus accumbens.

Importantly, obsessive-compulsive symptoms are not in general confined to primary OCD diagnoses but also may manifest across a spectrum of other neuropsychiatric disorders, such as TS^9^. This raises the possibility of a shared circuit-level substrate underpinning obsessive-compulsive behaviors that extends beyond diagnostic boundaries ^4,10^. To test this hypothesis, we examined a distinct cohort of patients with TS and comorbid obsessive-compulsive behavior who had undergone DBS targeting of either the amGPi or the cmThal. Overlap between E-fields and the OCD response tract (as defined by Li et al.^36^, based on data of patients with OCD), indeed, accounted for clinical improvements along the Y-BOCS in this TS cohort (R = 0.49, p = 0.019; **Fig. 4**). These findings suggest that modulating this circuit may be able to alleviate obsessive-compulsive symptomatology, regardless of diagnostic classification and stereotactic target.

**Fig. 4.**
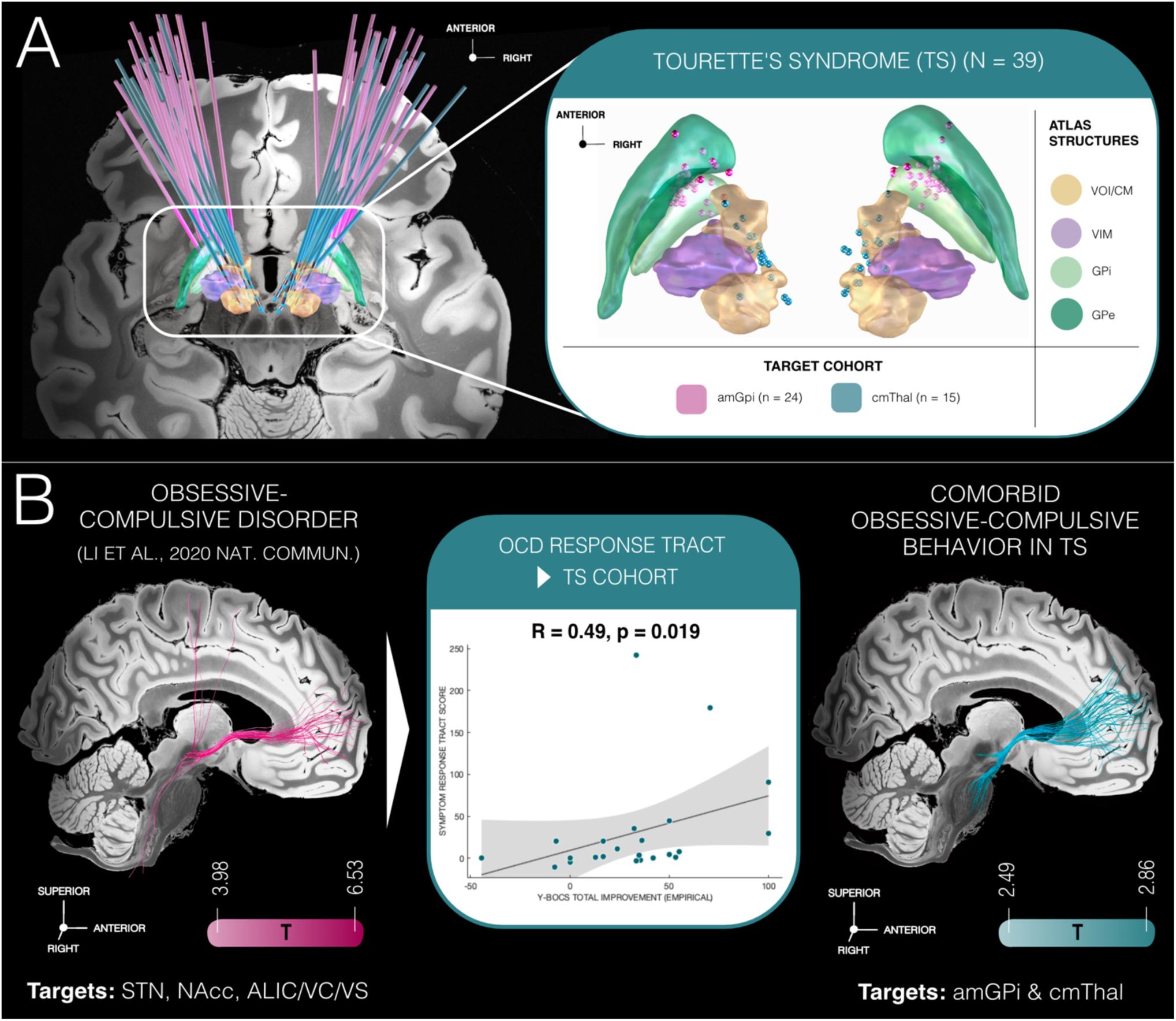
Relevance of obsessive-compulsive behavior circuitry irrespective of primary disorder. **(A)** Electrode placements of N = 39 patients with Tourette’s syndrome (TS) and comorbid obsessive-compulsive behavior. Each patient received bilateral electrodes to the anteromedial internal globus pallidus (amGPi, n = 24) or centromedian thalamic region (cmThal, n = 15). Electrodes are represented against a slice (z = -11 mm) from the 7T ex-vivo human brain atlas^63^ (*left*). The close-up (*right*) shows active electrode contacts, color-coded by target, in anatomical context of the ventrooralis internus/centromedian (VOI/CM) and ventral intermediate (VIM) nuclei of the thalamus, the external (GPe) and internal pallidum (GPi), extracted from the DBS Intrinsic Template atlas^60^, version 1.1. **(B)** The published obsessive-compulsive disorder (OCD) response tract^36^ (*left*) resembles that for comorbid obsessive-compulsive behavior for these TS patients (*right*), both plotted against a sagittal atlas^63^ slice (x = -5 mm). Each tract is color-coded by a T-value (consistent to Li et al.^36^), indicating its ability to distinguish good from poor responders. Overlap of TS patients’ E-fields with the tract model (resulting in one bilaterally averaged Symptom Response Tract Score per patient) correlates with Yale-Brown Obsessive-Compulsive Scale (Y-BOCS) outcomes (*middle*). These findings suggest a potential relevance of the tract, irrespective of primary disorder. Gray-shaded area indicates the 95% confidence interval. *Abbreviations:* ALIC, anterior limb of the internal capsule; NAcc, nucleus accumbens; VC/VS, ventral capsule/ventral striatum.

### A Multi-Symptom Circuit Taxonomy of OCD

While the OCD response tract correlated with overall symptom improvement, the disorder’s phenotypic heterogeneity encompasses wider symptom dimensions and frequent comorbidities that may point to a more complex neural architecture. We therefore hypothesized that the global tract could be further decomposed into finer-grained, symptom-specific response tracts that may be superior in explaining variability in treatment outcomes.

To investigate this possibility, we mapped the DBS response profiles of the four most prevalent symptom domains in OCD (obsessions, compulsions, depression, and anxiety) into anatomical tract space. Distinct yet overlapping symptom response tract profiles emerged, with hemispheric differences most prominently expressed for depression (**Fig. 5A**). Importantly, a highly similar topography of symptom mappings emerged both in the subset of patients with a complete set of data across all four core symptoms (n = 32; **Fig. 5A**) and in the full sample (N = 77; **Fig. S1A, Fig. 6).** This consistency strengthened our confidence that differential dysfunction attributions across symptoms were not simply driven by varying combinations of available symptom assessment data and stimulation targets.

**Fig. 5.**
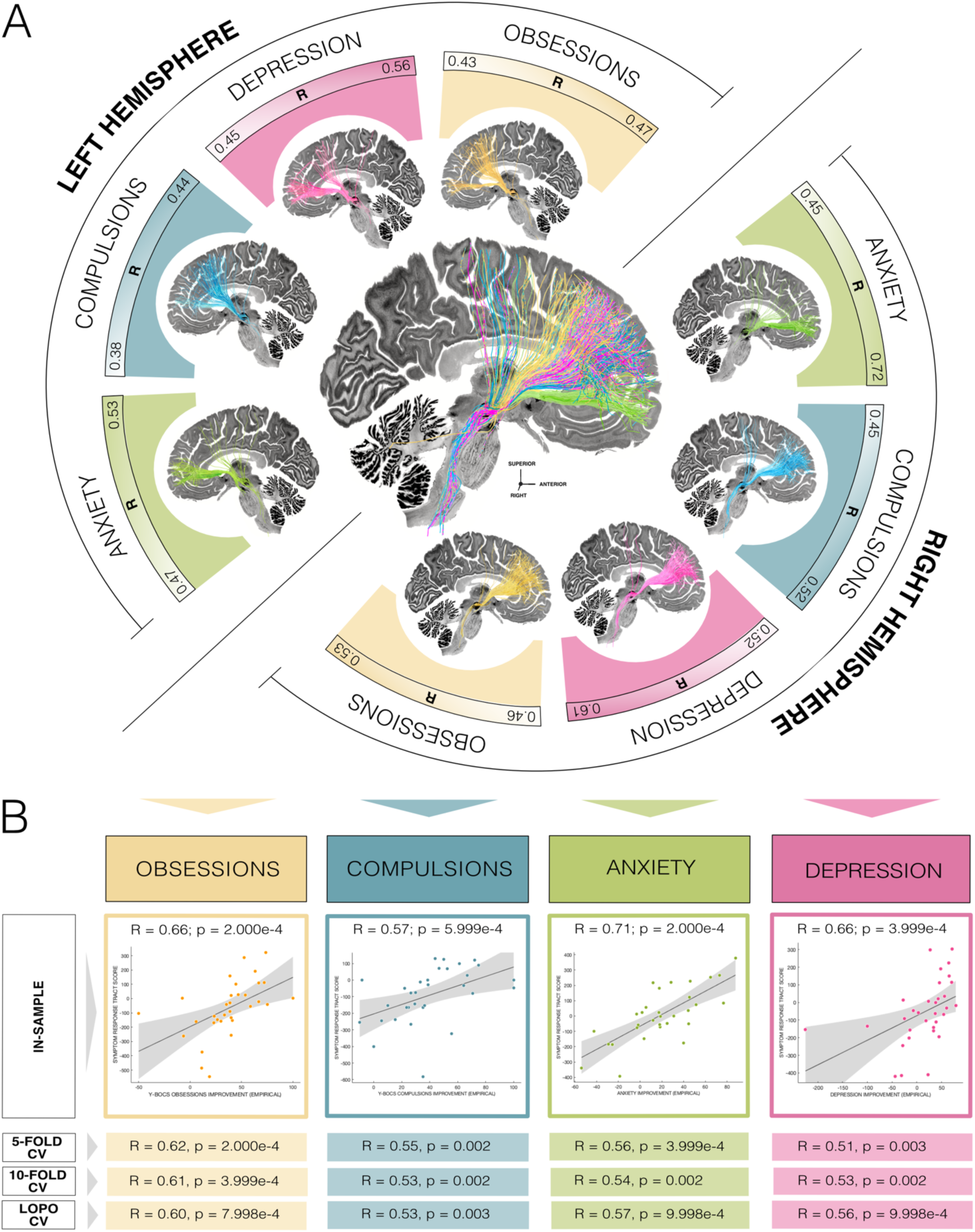
Multi-symptom taxonomy of deep brain stimulation (DBS) response tracts in obsessive-compulsive disorder (OCD). **(A)** The therapeutic response to DBS parcellates the brain’s connectome into a set of distinct yet partially overlapping symptom response tracts that are beneficial for alleviating obsessions, compulsions, anxiety, and depression in OCD. DBS Fiber Filtering results were derived from a high-resolution population-based connectome^66^ across 985 Human Connectome Project participants^67^ and superimposed on top of a sagittal slice (x = -5 mm) of the 100 µm BigBrain template^59^. Thresholded tract landscapes are represented in symptom-specific color, with color intensity graded by an R-value denoting the importance of modulating the respective tract for symptom benefit. Mappings were derived from the OCD subcohort (n = 32) with complete scores across all four core symptom domains, eliminating bias from variation in electrode placement across symptoms. Notably, results are consistent with those obtained from the full sample (N = 77; see Supplementary Materials). **(B)** Symptom response tracts were probed for their capability of explaining outcome variance within the sample (in-sample correlation). The Spearman correlation plots illustrate the association between degree of tract modulation (assessed by the Symptom Response Tract Score, averaged across bilateral estimates per patient) and empirical symptom outcome across the patient cohort. Additionally, various hold-out strategies – 5- and 10-fold, as well as leave-one-patient-out (LOPO) cross-validation (CV) – were performed, constructing connectivity models on subsets of the sample and estimating outcomes in held-out patients to allow for unbiased estimates. Gray shaded areas represent 95% confidence intervals.

**Fig. 6.**
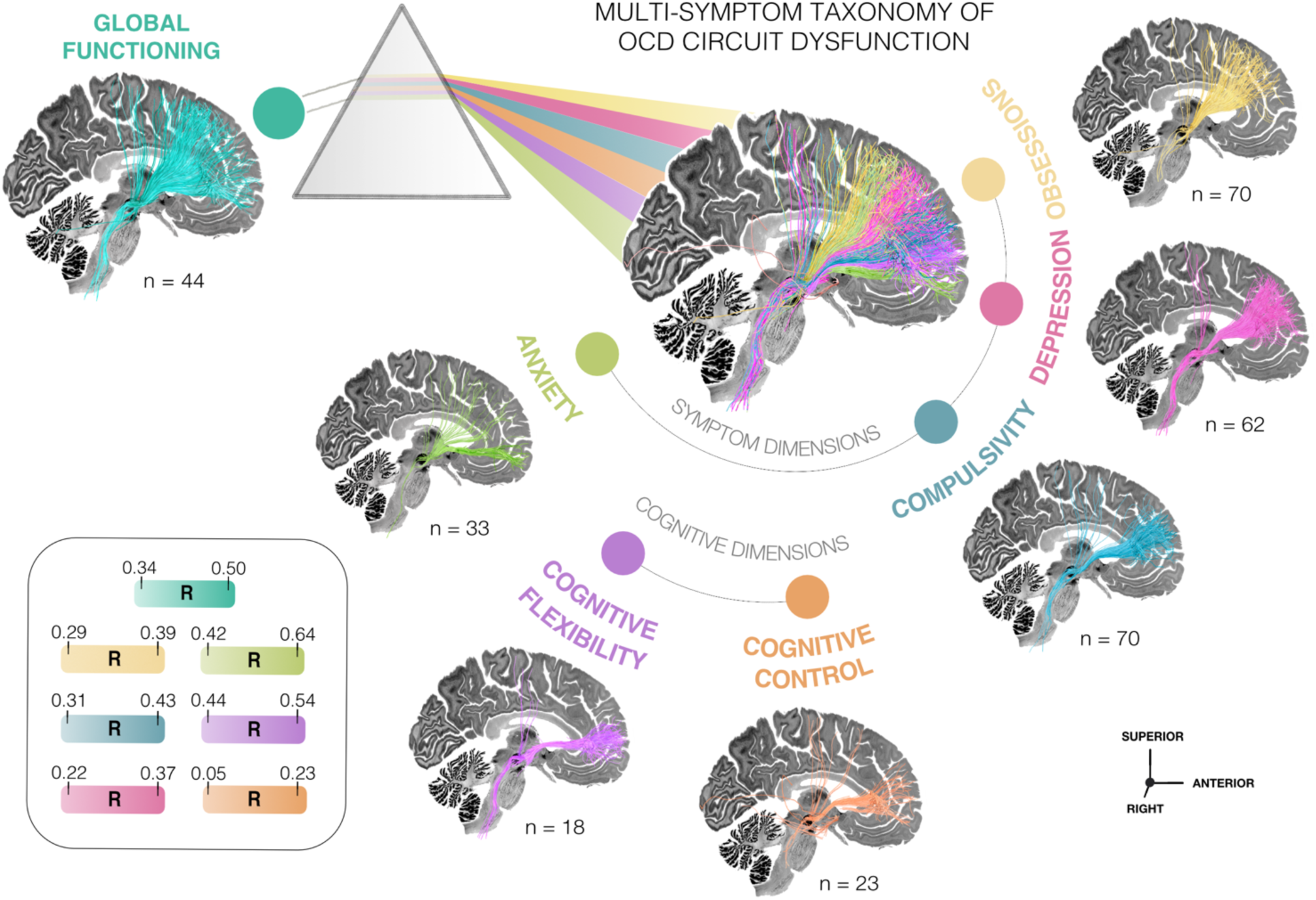
Multi-symptom foundation of everyday functioning. Global functioning and socio-occupational engagement benefit maps to a broad tract landscape within the internal capsule. By contrast, improvements in individual obsessive-compulsive disorder (OCD) symptoms (obsessions, compulsions, depression, anxiety, and deficits in cognitive control or flexibility) map to more constrained circuit subdomains of the anterior limb of the internal capsule. DBS Fiber Filtering results are informed on a high-resolution group connectome^66^ calculated based on data of 985 participants from the Human Connectome Project^67^, and displayed in thresholded form against a sagittal slice (x = -5 mm) of the 100 μm BigBrain template^59^. Each tract in the connectome is tagged with an R-value per symptom domain, reflecting the association between tract modulation and symptom outcomes across the cohort, with darker colors indicating stronger associations.

Anatomically, tracts associated with improvements in obsessive symptoms connected dorsolateral (dlPFC) and dorsomedial prefrontal (dmPFC), as well as ACC and pre-supplementary motor areas (pre-SMA) to subcortical stimulation sites, whereas compulsions mapped to connections with ACC, ventrolateral prefrontal cortices (vlPFC) and inferior frontal gyrus (IFG). The depression response tract, by contrast, exhibited higher asymmetry between hemispheres, with a connectivity profile involving dlPFC, ACC, and vlPFC in the right hemisphere. In the left hemisphere, the fibers were most strongly connected to the ventromedial prefrontal cortex (vmPFC) including OFC. A second left-hemispheric tract arm involved the dlPFC. The anxiety response tract predominantly projected from vmPFC with OFC but also comprised a secondary arm connecting stereotactic sites with dlPFC and pre-SMA. **Figs. S2** and **S3** provide axial and sagittal views of each tract to depict their anatomical features in greater detail.

**Fig. S4** shows negative response tracts, which were largely similar across symptoms. Critically, these tracts did not identify connections that actively worsened symptoms but were more likely driven by DBS cases in which optimal tracts were *not* stimulated based on the location of the implanted DBS lead. Presumably, any tracts that falls outside of the optimal set of connections will be tagged as negative due to the statistics applied in our analysis. Therefore, negative tracts may reflect suboptimal electrode placement rather than harmful connections. We draw this conclusion because only 4.3% of the patients with available scores showed symptom *worsening* in obsessions, 2.9% in compulsions, 23.8% in depression and 30.3% in anxiety.

When subjecting tract models to 5-fold, 10-fold and leave-one-patient-out (LOPO) cross-validation (CV), designed to prevent circularity bias in model-based estimates (see methods), their respective estimates significantly correlated with improvements in all four core symptoms (p < 0.05) in the subcohort of patients with a full set of complete scores. **Fig. 5B** represents an overview of all in-sample and CV results. Comparable results were reproduced in the full (N = 77) patient sample (**Fig. S1B**).

These four core symptom response tracts represent the principal results of our study. To expand upon these insights, we explored dysfunction in two cognitive domains that have demonstrated responsiveness to DBS in the context of OCD^52,64,65^, encompassing cognitive control and cognitive flexibility (**Fig. 6**). While these findings should be interpreted as preliminary due to the limited sample size, they may offer initial hypotheses for future investigation. **Fig. S5** shows the anatomical course of the exploratory tracts across different axial and sagittal 2D slices.

The cognitive control response tract (anchored to improvements in the Stroop test) was characterized by projections from the ACC, whereas the one for cognitive flexibility resembled the lower arm of the anxiety tract, with the most important involved cortical circuits projecting from vmPFC including OFC regions. Model validation yielded significant results for (circular) in-sample correlations (cognitive control: R = 0.47, p = 0.027; cognitive flexibility: R = 0.58, p = 0.013), but not for 5-fold (cognitive control: R = 0.28, p = 0.190; cognitive flexibility: R = -0.01, p = 0.961), 10-fold (cognitive control: R = 0.06, p = 0.696; cognitive flexibility: R = 0.15, p = 0.550), nor LOPO CV (cognitive control: R = 0.06, p = 0.725; cognitive flexibility: R = 0.12, p = 0.621).

**Fig. S6** demonstrates cortical surface projections of all symptom-wise response tracts separately (middle) as well as in synergy with each other.

### Multi-Symptom Response Tract Prediction of Global Functioning

Overall functioning in daily life and socio-occupational engagement, captured by the Global Assessment of Functioning (GAF), represents a broad, symptom-nonspecific construct that likely integrates contributions from multiple symptom domains that vary in salience across individuals. For example, in one patient severe anxiety may disproportionately drive GAF impairment, whereas in another, prominent compulsions may represent the dominant determinant. Based on the results presented above, we hypothesized that the GAF would map to the entire domain of the internal capsule in a less specific manner than in individual symptoms. This hypothesis was confirmed by our findings (**Fig. 6**, left).

We further reasoned that overall functional recovery in OCD following DBS may reflect the integrated modulation of multiple symptom-relevant circuits, rather than a single pathway in isolation. To test this, we calculated each patient’s mean predicted improvement across all six symptom-response tracts and compared its correlation with observed GAF changes to that of the GAF-specific tract alone. Stimulation volume overlaps with an exclusive GAF tract model (5-fold CV: R = 0.36, p = 0.012; 10-fold CV: R = 0.32, p = 0.040; LOPO CV: R = 0.34, p = 0.028) were outperformed by those from the combined multi-symptom model in explaining variance in observed GAF changes (5-fold CV: R = 0.45, p = 0.002; 10-fold CV: R = 0.48, p = 7.998e-4; LOPO CV: R = 0.48, p = 7.998e-4). These results indicate that coordinated engagement of multiple detailed symptom response circuits provides a more accurate account of recovery in everyday functioning than stimulation of the isolated GAF-associated tract by itself.

## Discussion

Obsessive-compulsive disorder (OCD) remains a formidable therapeutic challenge that is complicated by the expressed heterogeneity of its symptoms. To establish a circuit-based foundation for more personalized interventions, our study investigated how distinct patterns of fiber tract modulation related to DBS response in a large multi-center cohort of 77 OCD and 39 TS patients (244 electrodes in total) across 13 surgical centers and eight stereotactic targets.

Four principal conclusions arise from our findings. First, we validated the canonical OCD response tract, originally described in 2019/2020^35,36^ and repeatedly corroborated in independent patient cohorts^26,39,41–44^ in the present, larger OCD cohort. Second, engagement of this tract accounted for improvements in obsessive–compulsive behaviors in TS patients who underwent DBS of the amGPi or cmThal, indicating that a common anatomical pathway may mediate therapeutic response across targets and diagnostic categories. Third, this global tract was complemented by distinct yet overlapping circuit subdomains, each linked to improvements of different symptoms when analyzed for higher granularity. We investigated obsessions, compulsions, depression, and anxiety, but also dysfunction in two exploratory cognitive domains (cognitive control and flexibility). These symptom response tracts followed a topographic organization within the ALIC, with cortical peaks arranged along a dorsoventral axis and projecting from diverse prefrontal regions through the central ALIC toward striatum, STN, and brainstem. These pathways highlight the need for clinicians to fine-tune DBS targeting to different circuit disruptions. Fourth, a multi-symptom model incorporating all symptom response tracts provided a more robust account of functional improvement in daily life than a single-tract model.

These findings carry several important implications for our understanding of OCD and its optimized treatment. First, they highlight the disorder’s complex, multi-dimensional nature and limitations of categorial diagnostic systems, which may in part neglect neurobiological consideration^68–70^. In contrast, neuroimaging advances have driven a shift toward circuit-based models that recognize dysfunctions across multiple brain circuits as potential contributors to diversity in the OCD symptomatic phenotype^2,4–6,12^. Consistent with this notion, our distinct, yet overlapping, symptom response tracts challenge the traditional view that one selective anatomical target may address the entire, broad spectrum of OCD symptoms *in each individual patient*. Beyond the unifying response tract tied to global OCD benefit^46,47^, our results suggest the need for targeting (potentially interconnected) additional circuits. Each of these targets should modulate distinct symptom dimensions and be aimed at optimizing treatment outcomes. Concurrently, a partially shared recruitment of prefrontal regions across emotional, cognitive, and motor circuits will be integral to the understanding of OCD pathophysiology^2,4–6,12^, emphasizing that dysfunctions within these circuits do not operate in isolation, but along an interdependent network.

Second, the relationship between obsessions and compulsions remains actively debated^71^. Although OCD can be diagnosed in absence of one or the other, the two symptoms usually co-occur. The Y-BOCS provides separate subscales for both, acknowledging that they may function both independently and in tandem^72^. Classical cognitive–behavioral models that still influence psychotherapeutic practice cast compulsions as negatively reinforced avoidance responses to obsession-induced anxiety, implying a sequential or causal chain^73^. More recent accounts instead propose that compulsions could arise from a shift from goal-directed to habitual control^74,75^. Obsessions, in this view, may emerge secondarily as post-hoc justifications for actions that were otherwise difficult to rationalize, although this hypothesis remains untested^76^. Other perspectives see obsessions and compulsions as independent, yet co-occurring, cognitive and neural processes^77^. Our findings contribute to this debate by showing that obsessions and compulsions appear to recruit overlapping yet differentially weighted circuits. Their shared substrates align with the strongly correlated symptom improvements, whereas subtle circuit divergences suggest that optimal DBS targeting may differ slightly depending on whether obsessions or compulsions are prioritized. Programming strategies that preferentially activate contacts whose modeled stimulation fields overlap strongly with the targeted symptom response circuit could operationalize this distinction.

One possible explanation for the concurrent emergence of both symptoms is offered by the conflict processing theory of OCD, which emphasizes dysfunction in the dorsal ACC (dACC), a region implicated in both tract targets. The dACC is a region highly interconnected with different prefrontal, parietal, and motor regions, and among other functions, it acts as a gateway for cognitive and sensory information, intervening when conflicting information or incompatible response tendencies arise^78–80^. To resolve these conflicts, the ACC is thought to recruit the dlPFC, which then enhances attention to goal-related stimuli and assigns cognitive control. In OCD, dACC activation in response to internal or external conflicts fails to normalize, possibly due to ongoing OFC or striatal activation, creating a cycle of maladaptive conflict processing that impairs cognitive control^78^. By restoring balance in this circuitry, DBS may enhance both cognitive and emotional regulation and thus ease both obsessions and compulsions^78^. As such, DBS may mimic the effects of anterior cingulotomy, an ablative neurosurgical procedure targeting the ACC, or those of capsulotomy, targeting its subcortical projections^18,19,81^. Both the dACC (dmPFC) and dlPFC have further demonstrated effectiveness as noninvasive neuromodulation targets for OCD^82^.

In addition to shared substrates, the DBS response circuits for obsessions and compulsions exhibited *distinct* preferential mappings. Obsessive symptoms improved under DBS were linked to the most dorsal prefronto-subcortical projections, with peaks primarily emerging from pre-SMA, dlPFC, dmPFC, and ACC. This information closely matches the neuroimaging findings of divergent activity in these regions in OCD, with both hyper- and hypoactivation reported across study and task designs^83–90^. Implicated brain areas have been described as essential hubs within a dorsal cognitive circuit tasked with maintaining diverse executive functions, including working memory, attention, and planning for goal-directed decision-making and behavior, and its impairment is likely a deficit that contributes to various OCD symptoms^91,92^. In the context of obsessions, reduced cognitive control due to disrupted dlPFC activation may hinder the inhibition of intrusive thoughts and the management of associated emotional distress, reinforcing the cycle of obsessional thinking^7,92^. dlPFC dysfunction might also impair cognitive flexibility, rendering it difficult to shift attention away from obsessional thoughts^93^. Consistent with this hypothesis, noninvasive neuromodulation, including repetitive transcranial magnetic (rTMS) or direct current stimulation (tDCS) targeted to cortical hubs within the dorsal cognitive circuit, encompassing the dlPFC or pre-SMA, reduces OCD symptoms^82^.

These findings complement the identified compulsions response tract, which exhibited preferential subcortical connectivity with prefrontal regions including the ACC, and more ventral areas such as the vlPFC and IFG. These data suggest that modulating these connections plays a key role in inhibiting maladaptive rituals and restoring goal-directed behavior. Together with subcortical areas like the ventral caudate and thalamus, the IFG and vlPFC form a ventral cognitive circuit critical for self-regulation^2,5^. Neuroimaging studies also emphasize the importance of the IFG and particularly its subthalamic connections in response inhibition^94,95^. In OCD, individuals typically recognize the irrationality of their recurring thoughts and preemptive actions, which derails them from their intended goals. Deficits in inhibiting these unwanted behaviors may contribute to symptom persistence in OCD^7,77^, although some studies report conflicting results^96–98^. Relatedly, DBS treatments of OCD may restore impaired response inhibition, as signaled by enhanced theta oscillations in the IFG, alongside other regions, following DBS to the STN^99^ or VC/VS region^27,64^. Conflicting reports, however, have described *decreased* theta dynamics, as well^100,101^.

Third, once classified as an anxiety disorder, OCD shares marked overlap in its neuropsychological profile with this diagnostic category, particularly in dysregulated emotional responses to perceived threats^102,103^. While anxiety remains a core OCD feature, its precise role, whether as a primary symptom or an exacerbating factor for pre-existing obsessions and compulsions, remains to be clarified^5,12^. Our study identified an anxiety response tract that aligned with regions implicated in mood and anxiety disorders, particularly within the fronto-limbic network^104^, and it revealed peak activation in areas partially distinct from those associated with obsessions and compulsions. This network encompassed critical structures involved in generation (amygdala, NAcc) and evaluation (vmPFC, ventral OFC) of emotional responses, with a regulatory role of the dorsal cognitive circuit (dlPFC, dmPFC) at the nexus of cognitive and emotional processing^91^. In OCD, heightened limbic fear responses combined with impaired dorsal prefrontal top-down control may amplify anxiety by disrupting threat evaluation and regulation of emotional responses^13^. Impaired extinction learning of conditioned fear responses, potentially influenced by the vmPFC’s role in signaling safety after removal of a threat, may contribute to sustained anxiety^105,106^. Notably, effective DBS for OCD targeting key regions within this network, such as the NAcc, normalized aberrant fronto-limbic functional connectivity with the dm/dlPFC^107^.

Fourth, whether the depressive symptoms that frequently accompany OCD reflect secondary reactions to OCD-related distress or a comorbid circuit dysfunction remains unresolved^108^. The moderate correlation between improvements in depression and obsessions/compulsions in our cohort argues against depression being merely epiphenomenal. Consistently, the optimal DBS response tract for depression diverged from those for obsessions and compulsions, suggesting (partially) distinct mechanisms. Depression improvements mapped to right-hemispheric dlPFC, ACC, and vlPFC, and to left-hemispheric ventral prefrontal regions including vmPFC/OFC. The vmPFC, with the OFC, is central to the ventral affective circuit governing reward processing^2,5^. This circuit, in conjunction with the ventral striatum (particularly the NAcc), thalamus, and ACC, is not only commonly dysregulated in mood and anxiety disorders^104,109,110^, but also in OCD^105,111^. This is coherent with reduced reward sensitivity and exacerbated valuation of punishments observed in OCD^112,113^, which can contribute to anhedonia and negative emotional bias^114^. In OCD, compulsions may further provide relief from obsession-induced anxiety, creating a form of negative reinforcement that dampens the salience of natural rewards^112^. Consistent with a reward-circuit account, OFC hyperactivity reliably normalizes following effective OCD treatments, including pharmacotherapy and DBS^12^. Meanwhile, dysfunction in (cognitive) dlPFC-ACC circuits could contribute to impaired distress regulation^5,7,92^.

Ultimately, expanding the herein applied mapping strategy to a broader symptom range could contribute to an extensive taxonomy of the diverse circuit dysfunctions present in OCD, or the OCD “dysfunctome”, which may be defined by the set of symptom response tracts that can be selectively chosen, recombined, and modulated for treatment optimization ^39,115^. Beyond OCD, a similar approach may be applied to other disorders with comparably heterogenous presentations, that can be used to iteratively refine definitions of the human brain dysfunctome^39^ (**Fig. 7**). This concept aligns with the growing push for personalized neurosurgical interventions that are based on an individual’s unique symptom profile and neural circuitry^50,51,116–118^. Ongoing initiatives to identify transdiagnostic, dimensional symptoms such as compulsivity, anxiety, or reward learning highlight the critical overlaps between OCD and other psychiatric conditions, including addiction, TS, and impulse control disorders^5,10,11,119^. This hypothesis is reinforced by a shared response tract for obsessive-compulsive symptomatology in both DBS for OCD and TS, as demonstrated here and previously^44^. By uncovering unifying neurocognitive mechanisms, collaborative efforts like these could facilitate more flexible treatment strategies that extend beyond the traditional diagnostic boundaries of OCD^120^.

**Fig. 7.**
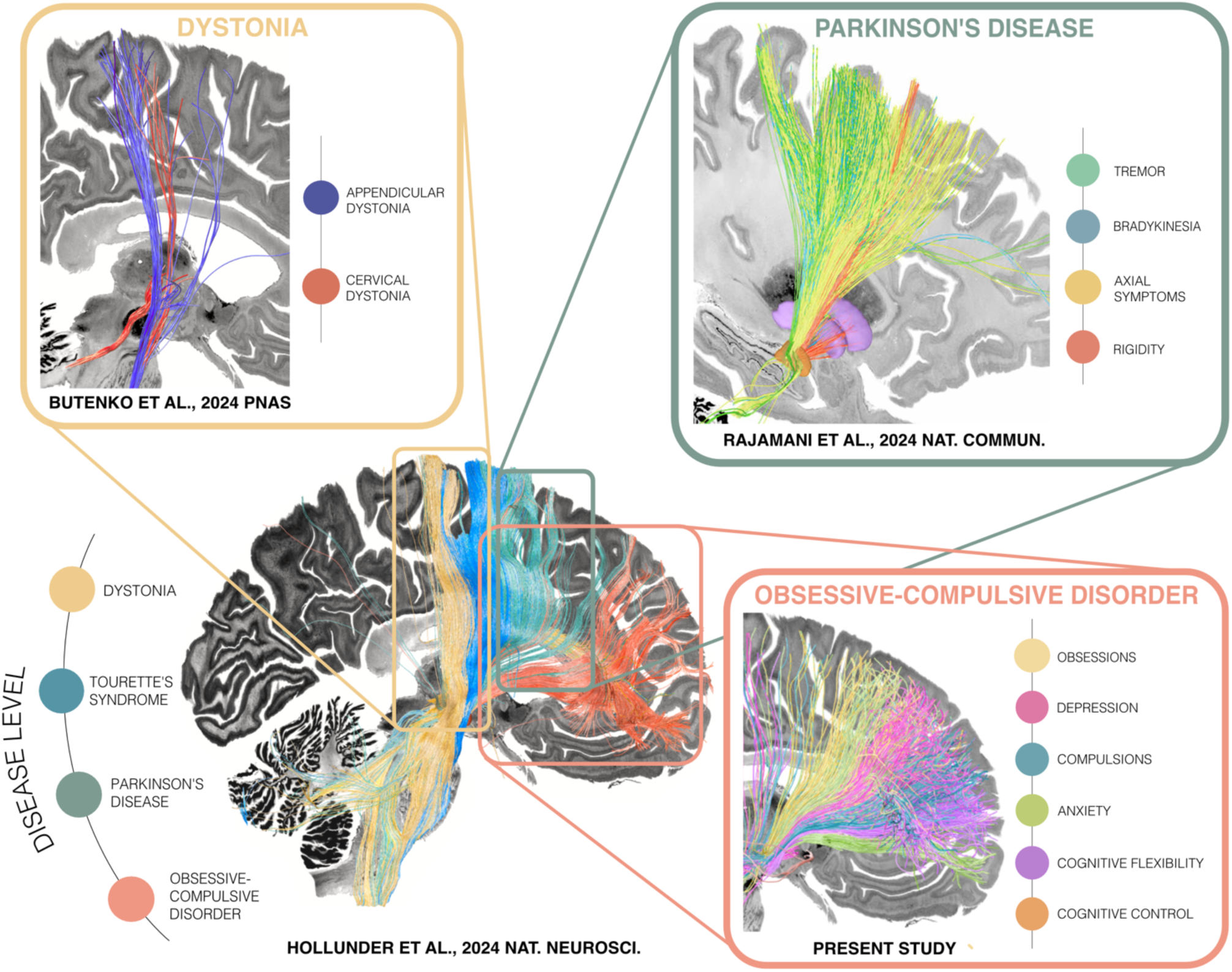
Mapping the circuit architecture of the human dysfunctome at increasing granularity. Hollunder et al. (2024)^39^ identified a shared fronto-subcortical axis along which deep brain stimulation (DBS) response tracts for dystonia, Tourette’s syndrome, Parkinson’s disease (PD), and obsessive-compulsive disorder (OCD) segregate. Building on this disease-level strategy, Rajamani et al. (2024)^51^ zeroed in on the PD motor response tract, uncovering distinct mappings for tremor, bradykinesia, rigidity, and axial symptom improvements. Butenko et al. (2025)^121^ applied the same principle to dystonia, distinguishing limb-related from axial symptom networks. The present study extends this trajectory by refining the OCD mapping, identifying detailed characterizations of connectomic topographies for OCD subdomains including obsessions, compulsions, depression, anxiety, cognitive control and cognitive flexibility. This layered approach — from broad diagnostic categories to more nuanced symptoms — may pave the way for increasingly flexible and personalized neuromodulation strategies with transdiagnostic potential. In the long term, applying this framework across diverse brain circuit disorders at increasingly detailed anatomical resolution may contribute to a comprehensive map of diverse circuit dysfunctions, or the human “dysfunctome”^39,115^.

Several limitations should be considered when interpreting our findings. First, the use of retrospective data may introduce bias in evaluating clinical outcomes. Incomplete assessments further resulted in differentially powered symptom response tracts, and pooling data from multiple centers may have introduced confounds, such as differences in clinical practice, patient selection, and data collection. However, the diversity of the large multi-center sample may have mitigated some of these factors, enhancing the study’s generalizability to real-world clinical conditions. Nonetheless, future prospective studies comprising standardized data collection should address these potential sources of bias.

Second, our primary circuit models were based on widely used, standardized rating instruments, either clinician-administered or self-reported. This approach supports clinical relevance and facilitates comparability across studies, which is essential for translational impact. Nonetheless, such measures carry inherent limitations, including subjectivity, sensitivity to symptom fluctuations and contextual factors. They may also not fully capture underlying neuropsychological mechanisms such as avoidance learning, habit formation, or response inhibition that could account for additional variability in circuit–phenotype relationships^74,75,77^. Future work incorporating both clinical measures and task-based or computational assessments may help address this gap.

Third, the E-field model employed in this study draws from a simplified representation of tissue activation and may therefore not fully reflect the underlying biophysical complexity^122^. Moreover, variations in stimulation parameters, such as pulse frequency or width, were neglected here yet may alter the interaction between stimulation and neural tissue^123^. Developments towards a more comprehensive model that takes additional factors into considerations may provide an increasingly detailed understanding of how stimulation influences brain activity^124^.

Fourth, the process of warping lead localizations into a template space may have introduced slight mismatches due to individual anatomical differences, which could affect the precision of reconstructed electrode placements and, consequently, the interpretation of stimulation effects. To minimize these errors, we employed advanced processing techniques, including brain shift correction^125^, multi-spectral normalization, and phantom-validated electrode reconstructions. Despite these efforts, some degree of error ultimately persists, and further refinement of lead localization techniques may help enhance the reliability of these mappings.

Finally, the use of normative connectivity data, due to limited availability of patient-specific diffusion-weighted MRI (dMRI) across included cohorts, may reduce the anatomical accuracy of mappings. Normative connectomes, while useful for generalizing across populations, do not capture individual variations found in brain connectivity or white-matter integrity that may be essential for precise surgical targeting or stimulation optimization. At the same time, normative connectivity is typically derived from datasets with higher resolution and superior signal-to-noise ratios than what can be acquired in clinical populations, owing to factors such as longer scan times and advanced acquisition protocols. A substantial body of work has also demonstrated the feasibility of robustly estimating clinical benefit from DBS based on normative connectivity information (for a review, see ^126^). Still, further validation of the herein identified circuit models with patient-specific data will represent essential complementary evidence to confirm their clinical translatability. Together, these findings may deepen our comprehension of the intricate circuit dysfunctions underlying OCD and the neural mechanisms through which DBS exerts its therapeutic effects. Beyond a global tract target associated with broad outcomes in obsessive-compulsive behavior irrespective of nominal diagnosis (OCD vs. TS) or stereotactic target, our study offers insights into the neural circuits underlying DBS response to more granular OCD symptoms. This may pave the way for increasingly nuanced treatment approaches tailored to the unique symptom constellations of each individual patient. This holistic perspective has the potential to enhance precision and efficacy of DBS for treatment-resistant OCD, while also offering broader implications for related neuropsychiatric disorders and other neuromodulation techniques.

## Methods and Materials

### Patient Cohorts, Imaging, and Clinical Assessments

#### OCD cohort

Connectivity models were calculated based on retrospective data from N = 77 multi-centric DBS patients, each treated for refractory OCD. Specifically, it comprised eight patient cohorts operated at the following institutions: University Hospital Cologne; Massachusetts General Hospital, Boston; Toronto Western Hospital; Hospital Clínico San Carlos, Madrid; National Hospital for Neurology and Neurosurgery, London; St Andrew’s War Memorial Hospital, Brisbane; St. Vincent’s Hospital Melbourne; and University Hospital Grenoble. Patients received bilateral DBS electrode implantation (Medtronic model 3387, n = 18; model 3389, n = 55; model 3391, n = 7; or St. Jude model 6143 ANS, n = 3) to one of six different stereotactic targets, namely to the ALIC (n = 29), NAcc (n = 15), ITP (n = 5), BNST (n = 9), or the STN (n = 13). Of note, London patients underwent concurrent stimulation of both STN and VC/VS targets (n = 6, four electrodes per patient), and stimulation settings as well as clinical scores corresponding to the “optimized double-stimulation” phase in this cohort were implemented^52^.

All patients with high-quality imaging for electrode reconstruction and clinical outcome data were included in the sample. Due to the uniqueness of the invasive data and its limited availability, the sample size was not pre-determined using statistical methods, but largely exceeds that of comparable investigations into the network effects of DBS in OCD^26,35–37,39,41,42,44^. To evaluate the power of our available sample for the intended analyses, we conducted a sensitivity analysis using G*Power software, version 3.1.9.6^127,128^. This approach indicated that our analyses achieve a statistical power greater than 78% for detecting effect sizes that have been reported in prior studies with comparable cross-validation designs concerning optimal connectivity profiles for DBS in movement disorders (R = 0.27–0.57) and neuropsychiatric conditions (R = 0.32–0.65)^126^, with an alpha level of 0.05 for one-tailed tests.

Preoperatively, magnetic resonance imaging (MRI) was performed in all patients for surgical planning. Postoperatively computed tomography (CT; n = 56) or MRI (n = 21) was implemented to verify the precise positioning of electrodes. Further demographic and clinical details on the analyzed cohorts are featured in **Table S1**. More extensive information on patient selection criteria, imaging parameters, surgical and programming procedures are provided in the respective original publications^26,29,35,52–55,149^.

Pre- to postoperative improvement was measured via continuous ratings on well-validated scales for obsessions vs. compulsions: Y-BOCS^72^; depression: Beck Depression Inventory (BDI)^129^, Montgomery-Åsperg Depression Rating Scale (MADRS)^130^, or Hamilton Depression Inventory (HAM-D)^131^; anxiety: Hamilton (HAM-A)^132^, and Beck Anxiety Inventories (BAI)^133^, and state section of the State-Trait Anxiety Inventory (STAI)^134^. To assess overall functioning and socio-occupational engagement, we further included the GAF^135^, given its broad availability across participating centers and continued use in clinical practice. While formal recommendation of the GAF for psychiatric assessment has been discontinued since 2013 due to limitations in inter-rater reliability and other psychometric properties^136,137^, its inclusion provides a pragmatic and widely interpretable measure. Any associated variability is unlikely to compromise the overall integrity of our findings.

To facilitate a standardized comparison despite the use of varying rating scales to measure similar constructs across cohorts, we expressed the respective scores as percentages of improvements from baseline as previously established in the literature^138–140^. Further, the number of available observations varied across the examined constructs (i.e., N = 77 for the Y-BOCS total score, n = 70 for both Y-BOCS obsessions and compulsions subscales, n = 63 for depression, n = 33 for anxiety, and n = 44 for global functioning; see **Table S1** for a detailed breakdown). To ensure consistent electrode placements across symptom response tract mappings, the main four-symptom tract model was derived from a subset of patients with complete data for all assessments (n = 32). Importantly, mappings were confirmed in the full cohort (N = 77). Because DBS effects in OCD often emerge gradually, all assessments were performed ≥3 months after stimulation activation to capture stabilized treatment effects.

In addition to these core symptom domains, we explored two cognitive features pooling available data of neuropsychiatric tests based on percentages of improvement from baseline to follow-up. First, cognitive control (total n = 23) was assessed by means of speed (n = 16) or number of correct answers within one minute (n = 7) in the incongruent condition of the Stroop Color-Word Test^141^, while cognitive flexibility (total n = 18) was evaluated based on log errors in the extra-dimensional stage (stage eight) of the Intra-Extra Dimensional Set Shifting subtask of the Cambridge Neuropsychological Test Automated Battery^142^ (n = 6) and time on the Trail Making Test (TMT)^143^ – part B (n = 12).

#### TS cohort

Finally, we validated the transdiagnostic relevance of the general OCD response tract target in a multi-center cohort of 39 TS patients with comorbid obsessive-compulsive behavior, pooled across seven cohorts from six contributing centers. Implanted with bilateral DBS electrodes in either the amGPi (n = 24) or the cmThal (n = 15), data of these patients were sourced from the International Tourette Syndrome DBS Database and Registry^58^ (https://tourettedeepbrainstimulationregistry.ese.ufhealth.org/) in collaboration with the International Neuromodulation Registry (https://neuromodulationregistry.org/). We incorporated all TS patients with high-quality imaging for electrode reconstruction and available Y-BOCS scores into our analyses. More granular obsessions and compulsions subscale data were unfortunately unavailable in a sufficiently large subcohort to permit meaningful analysis. Contributing centers comprised the IRCCS Istituto Ortopedico Galeazzi, Milan; University College London; University of Florida, Gainesville; New York University; Maastricht University Medical Center; and the University of California San Francisco. **Table S2** summarizes relevant imaging and electrode specifications for these cohorts along with demographic and clinical information.

All patients provided written informed consent for participation in the clinical trials and studies through their respective treatment centers. The current study, which involves secondary data analysis, received approval from Charité – Universitätsmedizin Berlin, and research procedures complied with the principles outlined in the Declaration of Helsinki throughout.

### DBS Electrode Localization and Electric Field Modeling

Preprocessing of patient imaging and electrode reconstruction were performed with aid of the Lead-DBS toolbox, v3.0^125,144,145^ (https://www.lead-dbs.org) which builds on MATLAB R2022b, v9.13.0.2105380 (The MathWorks Inc., Natick, MA, USA). The processing pipeline followed a repeatedly applied and well-established approach, which comprised linear coregistrations of postoperative CT or MRI anchored to preoperative T1-weighted images based on Advanced Normalization Tools^146^ (ANTs) (http://stnava.github.io/ANTs/), followed by correction for potential brain shift due to pneumocephalus during DBS surgery by means of an automated module. This step was ensued by multispectral spatial normalization of preoperative MRIs into template space (ICBM 2009b Non-linear Asymmetric) using the ANTs Symmetric Normalization (SyN) approach (“effective: low variance + subcortical refinement” preset within Lead-DBS). Atlas structures were defined by means of the DBS Intrinsic Template atlas (DISTAL)^60^, the California Institute of Technology reinforcement learning atlas (CIT168)^61^, or the Atlas of the Human Hypothalamus^62^ across analyzes and for visualization of results in anatomical context.

In patients with postoperative CT, electrodes were pre-localized using the Precise and Convenient Electrode Reconstruction for DBS (PaCER) algorithm^147^, while in patients with postoperative MRI, the trajectory search/contact reconstructions (TRAC/CORE) algorithm^125^ was implemented. Throughout, results underwent rigorous visual inspection by experienced users (B.H., G.M.M., and N.L., who are part of the Lead-DBS development team, with experience of more than 500 electrode reconstructions each), and manual adjustments via dedicated tools inherent to the Lead-DBS environment where appropriate.

Next, simulations of E-fields, which represent the gradient distribution of electrical potential in the patient’s native anatomical space, were performed using patient-specific stimulation parameters. This simulation was modeled using an adapted version of the SimBio/FieldTrip pipeline^148^ (available at https://www.mrt.uni-jena.de/simbio/ and http://fieldtriptoolbox.org/), as integrated into Lead-DBS. To create a volume conductor model, a finite element approach utilizing a four-compartment mesh was employed, which incorporates a detailed 3D representation of electrodes (metal and insulating aspects) and surrounding anatomical structures (gray and white matter). Subsequently, electrode trajectories and E-fields were transformed into template space. Group-level analysis was facilitated by the Lead-Group toolbox^149^ and the DBS Fiber Filtering Explorer^145^.

### Multi-Symptom Response Tract Modelling Procedure

Mapping symptoms to fiber tracts followed the established DBS Fiber Filtering approach^35^ in the version customized for non-binarized E-fields^150^, as described in detail in Hollunder et al. (2024)^39^ and repeatedly validated in prior work^35,39,43,51,150–152^. For comparability of mappings associated with DBS response in OCD to Li et al. (2020)^36^ and Hollunder et al. (2024)^39^, the foundation of our structural connectivity analysis consisted in a population-based group connectome^66^, derived from high-resolution dMRI tractography data of 985 healthy individuals obtained from the HCP 1,200 subjects release^67^. Specifics on this connectome are reported elsewhere^66^.

Following the strategy outlined in Hollunder et al. (2024)^39^, in brief, a “tract by E-field peak” matrix was first derived across all tracts in the connectome that were modulated at least moderately in a minimum fraction of the sample (i.e., traversed E-field magnitudes of at least 0.3 V/mm in more than 15% of patients), where each entry represented the maximum impact of each E-field on each tract. Next, weights were assigned to the activated tracts based on Spearman’s rank correlation between the matrix values with clinical improvements. Consequently, each tract was assigned an R-value, representing the relevance of its modulation for clinical outcome. The resulting tract profile can be viewed as a model of optimal connectivity for maximizing therapeutic benefit. Tracts with positive weights were those strongly influenced by E-fields in individuals who responded well to the treatment (referred to as “optimal symptom response tracts”), while tracts with negative weights were majorly influenced by E-fields in individuals who did not respond favorably to the treatment (“detrimental symptom response tracts”). This approach was applied for each symptom domain separately. Since correlation coefficients were determined through a mass-univariate approach, the accuracy of E-field overlap with these fiber profiles in estimating clinical outcome in novel data was validated (see below). **Fig. 1A** summarizes the methodological pipeline for this modeling stage.

Resulting symptom response tract models were visualized using Lead-DBS, v3.0 (for tract-based plots in 3D), MRIcroGL, v15.6.1 (https://www.nitrc.org/projects/mricrogl/) (for axial and sagittal 2D views of tracts), or Surf Ice software, v1.0.20211006 (https://www.nitrc.org/projects/surfice) (for surface-based plots). Axial and sagittal 2D plots were created by exporting the symptom response tract models in NIfTI format using Lead-DBS and smoothing them with a 1mm Gaussian kernel at full width at half maximum by means of SPM12 (https://www.fil.ion.ucl.ac.uk/spm/). Volumetric surface plots were created from volumetric density maps of exported tract models, after SPM-based smoothing with an 8 mm Gaussian kernel at full width at half maximum.

### Tract Model-Based Estimation of Symptom Outcomes

In accordance with published and repeatedly vetted methodological frameworks^51,115,150^, we then ran each of our symptom network models through a validation procedure in which their effectiveness in explaining pre- to postoperative symptom improvements in individual patients was probed (**Fig. 1B**). Again, this procedure was implemented for each symptom model independently. In brief, we first derived an estimate of the degree to which a patient’s E-fields modulated the optimal symptom response tract model. For each tract from the model covered by a given E-field, its model-based R-value was multiplied with the peak E-field magnitude. A mean across all these weighted R-values was subsequently calculated, so that each E-field was attributed with one corresponding “Symptom Response Tract Score”. Bilateral scores were finally averaged, and by that, each patient received one singular value.

The rationale behind this approach was as follows: E-field peaks that closely align with beneficial tracts are expected to yield high clinical scores, while those with little spatial alignment with beneficial or strong alignment with detrimental tracts would result in lower or negative clinical estimates. For this validation step, we selected the top 1,000 of both optimal and detrimental symptom response tracts based on the R-values of all tracts, thereby discarding less relevant ones for DBS-related symptom improvement from the model. To scrutinize the ability of fiber models to account for outcome variance within the sample, we correlated each patient’s Symptom Response Tract Score (as an estimate of clinical benefit) with their actual relative empirical symptom improvement following DBS in the respective symptom across the patient sample. This process ensured comparability of results across different symptoms.

To assess the ability of these models to explain clinical outcome variance in hold-out data (unbiased by circularity), we then subjected them to CV using two different k-fold (k = 5 and k = 10) as well as LOPO designs. In each fold, one fifth (5-fold CV), tenth (10-fold CV) or single patient (LOPO CV) was respectively held out of the sample as a test set, and the symptom response tract model was computed on the remaining patients (training set). Subsequently, Symptom Response Tract Scores were derived for the test set based on the model. Repeating this procedure across all k folds or patients (in the case of LOPO CV) resulted in one unbiased estimate per patient in the full sample. As a measure of goodness-of-fit, we finally calculated a Spearman correlation coefficient between Symptom Response Tract Scores and empirical improvements across the sample. One-sided tests were employed because CV tests focused exclusively on positive associations. Throughout, p-values were derived using permutation testing based on 5,000 iterations.

In addition, we intended to probe whether a predefined global symptom response tract target for DBS in OCD, as priorly published by Li et al. (2020)^35,36^, would also account for outcome in the present, more heterogeneous cohort. To this end, therapeutic benefit for each patient in the larger cohort was estimated by intersecting their E-fields with the published tract, using the same strategy described above. The resulting estimates were then correlated with empirical percentages of Y-BOCS improvements across the cohort to test for model validity in the present sample. To confirm the transdiagnostic validity of this global Y-BOCS response tract in TS data, we further applied a similar validation strategy in the N = 39 additional validation patients diagnosed with TS that exhibited comorbid obsessive-compulsive behavior. Again, symptom estimates were derived based on E-field overlap of validation patients with the Li et al. symptom response tract model^35,36^. Model fit was then assessed by comparing these Symptom Response Tract Scores with empirical Y-BOCS outcomes across the validation cohort using Spearman correlation.

Finally, we investigated whether a combined multi-symptom-based model would be able to more accurately estimate improvement in global level of functioning, as captured by the GAF, than the GAF response tract alone (single-tract model). To this end, one Symptom Response Tract Score was first derived for each patient based on each of the six symptom response tract models (obsessions, compulsions, anxiety, depression, cognitive flexibility, and cognitive control). The estimated improvement across the six symptom models was subsequently averaged and correlated with empirical outcome in the GAF. The performance of this composite symptom response tract score in estimating daily functioning was finally compared to that of the individual GAF response tract.

## Abbreviations

ACC: anterior cingulate cortex
ALIC: anterior limb of the internal capsule
amGPi: anteromedial internal pallidum
ANTs: Advanced Normalization Tools
BAI: Beck Anxiety Inventory
BDI: Beck Depression Inventory
BNST: bed nucleus of the stria terminalis
cmThal: centromedial thalamus
CIT168 atlas: California Institute of Technology reinforcement learning atlas
CV: Cross-validation
CT: computed tomography
DBS: deep brain stimulation
DISTAL atlas: DBS intrinsic template atlas
dACC: dorsal anterior cingulate cortex
dlPFC: dorsolateral prefrontal cortex
dMRI: diffusion-weighted magnetic resonance imaging
E-field: electric field magnitude
FEM: finite element method
GAF: Global Assessment of Functioning
GPe: external pallidum (globus pallidus, external segment)
GPi: internal pallidum (globus pallidus, internal segment)
HAM-A: Hamilton Anxiety Rating Scale
HAM-D: Hamilton Rating Scale for Depression
HCP: Human Connectome Project
IFG: inferior frontal gyrus
ITP: inferior thalamic peduncle
JHU atlas: Johns Hopkins University atlas
MADRS: Montgomery-Åsberg Depression Rating Scale
MGH: Massachusetts General Hospital
MNI space: ICBM 2009b Non-linear Asymmetric template space
MRI: magnetic resonance imaging
NAcc: nucleus accumbens
OCD: obsessive-compulsive disorder
OFC: orbitofrontal cortex
PaCER algorithm: Precise and Convenient Electrode Reconstruction for Deep Brain Stimulation algorithm
pre-SMA: pre-supplementary motor area
rTMS: repetitive transcranial magnetic stimulation
sl-MFB: superolateral branch of the medial forebrain bundle
STN: subthalamic nucleus
SyN approach: Symmetric Normalization approach
tDCS: transcranial direct current stimulation
TRAC/CORE algorithm: trajectory search/contact reconstructions algorithm
VC/VS: ventral capsule/ventral striatum
VIM: ventral intermediate nucleus of the thalamus,
vlPFC: ventrolateral prefrontal cortex
vmPFC: ventromedial prefrontal cortex
VOI/CM: nucleus ventrooralis internus/centromedian nucleus of the thalamus
Y-BOCS: Yale-Brown Obsessive-Compulsive Scale

## Data Availability

Due to data protection regulations, individual patient data cannot be made publicly available, as doing so would compromise privacy. However, these data may be obtained from the principal investigators at the respective collection sites upon reasonable request and within the framework of a data-sharing agreement. Requests for additional information or access to data should be directed to the corresponding author of this manuscript, B.H., who is committed to responding within a maximum of 30 days. Files and scripts to use the HCP 985 Connectome^66^ within the Lead-DBS software environment can be downloaded via the following link: https://www.lead-dbs.org/helpsupport/knowledge-base/atlasesresources/normative-connectomes/. Source data used for construction of this connectome are freely accessible through the Human Connectome Project repository: https://www.humanconnectome.org/study/hcp-young-adult/document/1200-subjects-data-release. The DISTAL atlas^60^, version 1.1, and the CIT168^61^ atlas, version 1.1, are both openly available via the Lead-DBS knowledge base (https://www.lead-dbs.org/helpsupport/knowledge-base/atlasesresources/atlases-2/) and are provided as pre-installations with the Lead-DBS software. The Atlas of the Human Hypothalamus^62^, version 1.0, is also made openly accessible by the original authors at: https://zenodo.org/records/3942115.

## Code Availability

All code used in the analyses presented in this study is openly accessible as part of the Lead-DBS software environment, available at: https://github.com/netstim/leaddbs.

## Acknowledgements

The authors would like to express their gratitude to the patients for their study participation. B.H. and

I.A.S. gratefully acknowledge support by a scholarship from the Einstein Center for Neurosciences Berlin. B.H. is also grateful for support by the Prof. Dr. Klaus Thiemann Foundation (Parkinson Fellowship 2024). A.H. was supported by the German Research Foundation (Deutsche Forschungsgemeinschaft, 424778381 – TRR 295), Deutsches Zentrum für Luft- und Raumfahrt (DynaSti grant within the EU Joint Programme Neurodegenerative Disease Research, JPND), the National Institutes of Health (R01MH130666, 1R01NS127892-01, 2R01 MH113929 & UM1NS132358) as well as the New Venture Fund (FFOR Seed Grant). The Department of Clinical and Movement Neurosciences, UCL Queen Square Institute of Neurology, London, UK is supported by the Medical Research Council, the National Institute for Health and Care Research, and the University College London Hospitals Biomedical Research Centre. J.C.B. was supported by the Else-Kröner-Fresenius-Stiftung (2022_EKES.23).

## Authorship Contribution Statement

B.H.: conceptualization, data curation, methodology, formal analysis, visualization, writing — original draft, and funding acquisition. G.M.M., N.L.: data curation, methodology, software, formal analysis, writing — review and editing. P.R.: methodology, formal analysis, visualization, writing — review and editing. I.A.S., J.P.: data curation, formal analysis, writing — review and editing. N.R.: methodology, software, resources, writing — review and editing. C.N., P.M., N.A., K.A.J., N.P., H.B., H.A., B.M.B., T.C., J.G., J.P.C.M., B.S., J.A.B., H.T., D.J.C., S.R., P.B., C.F., A.A.K., D.S., A.B., M.P., A.Y.M., M.H.P., J.L.O., J.K., V.V.-V., S.C., S.A.S, W.K.G., A.H.S., K.S.C., B.H.K., H.S.M., M.F., T.F., P.L., L.A., J.J.A.dJ., A.F.G.L., C.R.B., M.S.O., R.M.R., G.R.C., D.D.D., S.H.S., A.M.L., L.Z., E.J., M.P., J.C.B., M.D.F.: investigation, resources, data curation, writing — review and editing. C.N.: investigation, resources, data curation, formal analysis, writing — review and editing. A.H.: conceptualization, methodology, software, resources, formal analysis, visualization, writing — original draft, supervision, project administration, and funding acquisition.

## References

1. Stein, D. J. et al. Obsessive compulsive disorder. Nat Rev Dis Primers 5, 1–46 (2020).

2. Shephard, E. et al. Toward a neurocircuit-based taxonomy to guide treatment of obsessive–compulsive disorder. Molecular Psychiatry 26, 4583–4604 (2021).

3. Leckman, J. F. et al. Obsessive-compulsive disorder: A review of the diagnostic criteria and possible subtypes and dimensional specifiers for DSM-V. Depression and Anxiety 27, 507–527 (2010).

4. Robbins, T. W., Vaghi, M. M. & Banca, P. Obsessive-compulsive disorder: Puzzles and prospects. Neuron 102, 27–47 (2019).

5. van den Heuvel, O. A., et al. Brain circuitry of compulsivity. European Neuropsychopharmacology 26, 810–827 (2016).

6. Thorsen, A. L., Kvale, G., Hansen, B. & van den Heuvel, O. A. Symptom dimensions in obsessive-compulsive disorder as predictors of neurobiology and treatment response. Current Treatment Options in Psychiatry 5, 182–194 (2018).

7. Abramovitch, A., Abramowitz, J. S. & Mittelman, A. The neuropsychology of adult obsessive-compulsive disorder: A meta-analysis. Clinical Psychology Review 33, 1163–1171 (2013).

8. Eisen, J. L. et al. Impact of obsessive-compulsive disorder on quality of life. Comprehensive Psychiatry 47, 270–275 (2006).

9. Hirschtritt, M. E. et al. Lifetime prevalence, age of risk, and etiology of comorbid psychiatric disorders in Tourette syndrome. JAMA Psychiatry 72, 325–333 (2015).

10. Gillan, C. M., Fineberg, N. A. & Robbins, T. W. A trans-diagnostic perspective on obsessive-compulsive disorder. Psychological Medicine 47, 1528–1548 (2017).

11. Robbins, T. W., Banca, P. & Belin, D. From compulsivity to compulsion: the neural basis of compulsive disorders. Nature Reviews Neuroscience 25, 313–333 (2024).

12. Ahmari, S. E. & Rauch, S. L. The prefrontal cortex and OCD. Neuropsychopharmacology 47, 211–224 (2022).

13. Mataix-Cols, D. & van den Heuvel, O. A. Common and distinct neural correlates of obsessive-compulsive and related disorders. Psychiatric Clinics of North America 29, 391–410 (2006).

14. Raviv, N. et al. A systematic review of deep brain stimulation targets for obsessive compulsive disorder. Neurosurgery 87, 1098–1110 (2020).

15. Hirschtritt, M. E., Bloch, M. H. & Mathews, C. A. Obsessive-compulsive disorder advances in diagnosis and treatment. JAMA - Journal of the American Medical Association 317, 1358–1367 (2017).

16. Nuttin, B., Cosyns, P., Demeulemeester, H., Gybels, J. & Meyerson, B. Electrical stimulation in anterior limbs of internal capsules in patients with obsessive-compulsive disorder. The Lancet 354, 1526 (1999).

17. Kohl, S. et al. Deep brain stimulation for treatment-refractory obsessive compulsive disorder: A systematic review. BMC Psychiatry 14, 1–10 (2014).

18. Fodstad, H., Strandman, E., Karlsson, B. & West, K. A. Treatment of chronic obsessive compulsive states with stereotactic anterior capsulotomy or cingulotomy. Acta Neurochirurgica 62, 1–23 (1982).

19. Brown, L. T. et al. Dorsal anterior cingulotomy and anterior capsulotomy for severe, refractory obsessive-compulsive disorder: a systematic review of observational studies. Journal of Neurosurgery 124, 77–89 (2016).

20. Menchón, J. M. et al. A prospective international multi-center study on safety and efficacy of deep brain stimulation for resistant obsessive-compulsive disorder. Molecular Psychiatry 26, 1234–1247 (2021).

21. Denys, D. et al. Efficacy of deep brain stimulation of the ventral anterior limb of the internal capsule for refractory obsessive-compulsive disorder: A clinical cohort of 70 patients. American Journal of Psychiatry 177, 265–271 (2020).

22. Greenberg, B. D. et al. Deep brain stimulation of the ventral internal capsule/ventral striatum for obsessive-compulsive disorder: Worldwide experience. Molecular Psychiatry 15, 64–79 (2010).

23. Denys, D. et al. Deep brain stimulation of the nucleus accumbens for treatment-refractory obsessive-compulsive disorder. Archives of General Psychiatry 67, 1061–1068 (2010).

24. Sturm, V. et al. The nucleus accumbens: A target for deep brain stimulation in obsessive-compulsive- and anxiety-disorders. Journal of Chemical Neuroanatomy 26, 293–299 (2003).

25. Provenza, N. R. et al. Long-term ecological assessment of intracranial electrophysiology synchronized to behavioral markers in obsessive-compulsive disorder. Nat Med 27, 2154–2164 (2021).

26. Mosley, P. E. et al. A randomised, double-blind, sham-controlled trial of deep brain stimulation of the bed nucleus of the stria terminalis for treatment-resistant obsessive-compulsive disorder. Translational Psychiatry 11, 190 (2021).

27. Provenza, N. R. et al. Disruption of neural periodicity predicts clinical response after deep brain stimulation for obsessive-compulsive disorder. Nat Med 30, 3004–3014 (2024).

28. Shofty, B. et al. Intraoperative valence testing to adjudicate between ventral capsule/ventral striatum and bed nucleus of the stria terminalis target selection in deep brain stimulation for obsessive-compulsive disorder. Journal of Neurosurgery 139, 442–450 (2023).

29. Lee, D. J. et al. Inferior thalamic peduncle deep brain stimulation for treatment-refractory obsessive-compulsive disorder: A phase 1 pilot trial. Brain Stimulation 12, 344–352 (2019).

30. Chabardes, S. et al. Deep brain stimulation of the subthalamic nucleus in obsessive-compulsives disorders: Long-term follow-up of an open, prospective, observational cohort. Journal of Neurology, Neurosurgery and Psychiatry 91, 1349–1356 (2020).

31. Mallet, L. et al. Subthalamic nucleus stimulation in severe obsessive-compulsive disorder. New England Journal of Medicine 359, 2121–2134 (2008).

32. Coenen, V. A. et al. The medial forebrain bundle as a target for deep brain stimulation for obsessive-compulsive disorder. CNS Spectrums 22, 282–289 (2017).

33. Alonso, P. et al. Deep brain stimulation for obsessive-compulsive disorder: A meta-analysis of treatment outcome and predictors of response. PLoS ONE 10, e0133591 (2015).

34. Gadot, R. et al. Efficacy of deep brain stimulation for treatment-resistant obsessive-compulsive disorder: systematic review and meta-analysis. Journal of Neurology, Neurosurgery and Psychiatry 93, 1166–1173 (2022).

35. Baldermann, J. C. et al. Connectivity profile predictive of effective deep brain stimulation in obsessive-compulsive disorder. Biological Psychiatry 85, 735–743 (2019).

36. Li, N. et al. A unified connectomic target for deep brain stimulation in obsessive-compulsive disorder. Nature Communications 11, 3364 (2020).

37. Li, N. et al. A unified functional network target for deep brain stimulation in obsessive-compulsive disorder. Biological Psychiatry 90, 701–713 (2021).

38. Haber, S. N., Yendiki, A. & Jbabdi, S. Four Deep Brain Stimulation Targets for Obsessive-Compulsive Disorder: Are They Different? Biological Psychiatry 90, 667–677 (2021).

39. Hollunder, B. et al. Mapping dysfunctional circuits in the frontal cortex using deep brain stimulation. Nature Neuroscience 27, 573–586 (2024).

40. Horn, A. et al. Deep Brain Stimulation response circuits in Obsessive Compulsive Disorder. Biological Psychiatry S0006322325010960 (2025) doi:10.1016/j.biopsych.2025.03.008.

41. van der Vlis, T. A. M. B. et al. Ventral capsule/ventral striatum stimulation in obsessive-compulsive disorder: Toward a unified connectomic target for deep brain stimulation? Neuromodulation 24, 316–323 (2021).

42. Smith, A. H. et al. Replicable effects of deep brain stimulation for obsessive-compulsive disorder. Brain Stimulation 14, 1–3 (2021).

43. Gadot, R. et al. Tractography-based modeling explains treatment outcomes in patients undergoing deep brain stimulation for obsessive-compulsive disorder. 10.1016/j.biopsych.2023.01.017 doi:10.1016/j.biopsych.2023.01.017.

44. Johnson, K. A. et al. Basal ganglia pathways associated with therapeutic pallidal deep brain stimulation for Tourette syndrome. Biological Psychiatry: Cognitive Neuroscience and Neuroimaging 10.1016/j.bpsc.2020.11.005 (2020) doi:10.1016/j.bpsc.2020.11.005.

45. Coenen, V. A. et al. Deconstructing a common pathway concept for Deep Brain Stimulation in the case of Obsessive-Compulsive Disorder. Molecular Psychiatry 10.1038/s41380-025-03008-x (2025) doi:10.1038/s41380-025-03008-x.

46. Baldermann, J. C. et al. Connectomic deep brain stimulation for obsessive-compulsive disorder. Biological Psychiatry 90, 678–688 (2021).

47. Horn, A. et al. Deep brain stimulation response circuits in obsessive-compulsive disorder. Biological Psychiatry 10.1016/j.biopsych.2025.03.008 (2025) doi:10.1016/j.biopsych.2025.03.008.

48. Basich-Pease, G. et al. Tractography-based DBS lead repositioning improves outcome in refractory OCD and depression. Front. Hum. Neurosci. 17, 1339340 (2024).

49. Campbell, J. M. et al. Tractography-guided DBS programming in treatment-resistant OCD: A case report and review of the literature. Biological Psychiatry S0006322325015380 (2025) doi:10.1016/j.biopsych.2025.10.017.

50. Hollunder, B. et al. Toward personalized medicine in connectomic deep brain stimulation. Progress in Neurobiology 210, 102211 (2022).

51. Rajamani, N. et al. Deep brain stimulation of symptom-specific networks in Parkinson’s disease. Nature Communications 15, 1–16 (2024).

52. Tyagi, H. et al. A randomized trial directly comparing ventral capsule and anteromedial subthalamic nucleus stimulation in obsessive-compulsive disorder: Clinical and imaging evidence for dissociable effects. Biological Psychiatry 85, 726–734 (2019).

53. Barcia, J. A. et al. Personalized striatal targets for deep brain stimulation in obsessive-compulsive disorder. Brain Stimulation 12, 724–734 (2019).

54. Polosan, M. et al. Affective modulation of the associative-limbic subthalamic nucleus: deep brain stimulation in obsessive–compulsive disorder. Translational Psychiatry 9, (2019).

55. McLaughlin, N. C. R. et al. Double blind randomized controlled trial of deep brain stimulation for obsessive-compulsive disorder: Clinical trial design. Contemporary Clinical Trials Communications 22, 100785 (2021).

56. Kisely, S. et al. Deep brain stimulation for obsessive-compulsive disorder: A systematic review and meta-analysis. Psychological Medicine 44, 3533–3542 (2014).

57. Mar-Barrutia, L. et al. Deep brain stimulation for obsessive-compulsive disorder: A systematic review of worldwide experience after 20 years. World Journal of Psychiatry 11, 659–680 (2021).

58. Martinez-Ramirez, D. et al. Efficacy and safety of deep brain stimulation in tourette syndrome the international tourette syndrome deep brain stimulation public database and registry. JAMA Neurology 75, 353–359 (2018).

59. Amunts, K. et al. BigBrain: An ultrahigh-resolution 3D human brain model. Science 340, 1472–1475 (2013).

60. Ewert, S. et al. Toward defining deep brain stimulation targets in MNI space: A subcortical atlas based on multimodal MRI, histology and structural connectivity. NeuroImage 170, 271–282 (2018).

61. Pauli, W. M., Nili, A. N. & Tyszka, J. M. A high-resolution probabilistic in vivo atlas of human subcortical brain nuclei. Scientific Data 5, 180063 (2018).

62. Neudorfer, C. et al. A high-resolution in vivo magnetic resonance imaging atlas of the human hypothalamic region. Scientific Data 7, 1–14 (2020).

63. Edlow, B. L. et al. 7 Tesla MRI of the ex vivo human brain at 100 micron resolution. Scientific Data 6, 244 (2019).

64. Widge, A. S. et al. Deep brain stimulation of the internal capsule enhances human cognitive control and prefrontal cortex function. Nature Communications 10, 1–11 (2019).

65. Bouwens Van Der Vlis, T. A. M., et al. Cognitive outcome after deep brain stimulation for refractory obsessive–compulsive disorder: A systematic review. Neuromodulation: Technology at the Neural Interface 25, 185–194 (2022).

66. Elias, G. J. B. et al. A large normative connectome for exploring the tractographic correlates of focal brain interventions. Scientific Data 11, 1–12 (2024).

67. Van Essen, D. C. et al. The WU-Minn Human Connectome Project: An overview. NeuroImage 80, 62–79 (2013).

68. Insel, T. R. The NIMH Research Domain Criteria (RDoC) Project: Precision medicine for psychiatry. American Journal of Psychiatry 171, 395–397 (2014).

69. Cuthbert, B. N. The RDoC framework: Facilitating transition from ICD/DSM to dimensional approaches that integrate neuroscience and psychopathology. World Psychiatry 13, 28–35 (2014).

70. Insel, T. et al. Research Domain Criteria ( RDoC ): Toward a new classification framework for research on mental disorders. American Journal of Psychiatry 167, 748–751 (2010).

71. Kalanthroff, E. & Wheaton, M. G. An integrative model for understanding obsessive-compulsive disorder: Merging cognitive behavioral theory with insights from clinical neuroscience. Journal of Clinical Medicine 11, (2022).

72. Goodman, W. K. et al. The Yale-Brown Obsessive Compulsive Scale: I. Development, use, and reliability. Archives of General Psychiatry 46, 1006–1011 (1989).

73. Abramowitz, J. S., Taylor, S. & McKay, D. Obsessive-compulsive disorder. The Lancet 374, 491–499 (2009).

74. Gillan, C. M. et al. Disruption in the balance between goal-directed behavior and habit learning in obsessive-compulsive disorder. American Journal of Psychiatry 168, 718–726 (2011).

75. Voon, V. et al. Disorders of compulsivity: A common bias towards learning habits. Molecular Psychiatry 20, 345–352 (2015).

76. Gillan, C. M. & Sahakian, B. J. Which is the driver, the obsessions or the compulsions, in OCD? Neuropsychopharmacology 40, 247–248 (2015).

77. Chamberlain, S. R., Blackwell, A. D., Fineberg, N. A., Robbins, T. W. & Sahakian, B. J. The neuropsychology of obsessive compulsive disorder: The importance of failures in cognitive and behavioural inhibition as candidate endophenotypic markers. Neuroscience and Biobehavioral Reviews 29, 399–419 (2005).

78. McGovern, R. A. & Sheth, S. A. Role of the dorsal anterior cingulate cortex in obsessive-compulsive disorder: Converging evidence from cognitive neuroscience and psychiatric neurosurgery. Journal of Neurosurgery 126, 132–147 (2017).

79. Van Veen, V., Cohen, J. D., Botvinick, M. M., Stenger, V. A. & Carter, C. S. Anterior cingulate cortex, conflict monitoring, and levels of processing. NeuroImage 14, 1302–1308 (2001).

80. Braver, T. S., Barch, D. M., Gray, J. R., Molfese, D. L. & Snyder, A. Anterior cingulate cortex and response conflict: Effects of frequency, inhibition and errors. Cerebral Cortex 11, 825–836 (2001).

81. Greenberg, B. D., Rauch, S. L. & Haber, S. N. Invasive circuitry-based neurotherapeutics: Stereotactic ablation and deep brain stimulation for OCD. Neuropsychopharmacology 35, 317–336 (2010).

82. Fitzsimmons, S. M. D. D. et al. Repetitive transcranial magnetic stimulation for obsessive-compulsive disorder: A systematic review and pairwise/network meta-analysis. Journal of Affective Disorders 302, 302–312 (2022).

83. Adler, C. M. et al. fMRI of neuronal activation with symptom provocation in unmedicated patients with obsessive compulsive disorder. Journal of Psychiatric Research 34, 317–324 (2000).

84. Cheng, Y. et al. Abnormal resting-state activities and functional connectivities of the anterior and the posterior cortexes in medication-naïve patients with obsessive-compulsive disorder. PLoS ONE 8, (2013).

85. Fitzgerald, K. D. et al. Error-related hyperactivity of the anterior cingulate cortex in obsessive-compulsive disorder. Biological Psychiatry 57, 287–294 (2005).

86. Gruner, P. et al. Independent component analysis of resting state activity in pediatric obsessive-compulsive disorder. Human Brain Mapping 35, 5306–5315 (2014).

87. Maia, T. V., Cooney, R. E. & Peterson, B. S. The neural bases of obsessive - Compulsive disorder in children and adults. Development and Psychopathology 20, 1251–1283 (2008).

88. Maltby, N., Tolin, D. F., Worhunsky, P., O’Keefe, T. M. & Kiehl, K. A. Dysfunctional action monitoring hyperactivates frontal-striatal circuits in obsessive-compulsive disorder: An event-related fMRI study. NeuroImage 24, 495–503 (2005).

89. Thorsen, A. L. et al. Emotional processing in obsessive-compulsive disorder: A systematic review and meta-analysis of 25 functional neuroimaging studies. Biological Psychiatry: Cognitive Neuroscience and Neuroimaging 3, 563–571 (2018).

90. Schlösser, R. G. M. et al. Fronto-cingulate effective connectivity in obsessive compulsive disorder: A study with fMRI and dynamic causal modeling. Human Brain Mapping 31, 1834–1850 (2010).

91. Friedman, N. P. & Robbins, T. W. The role of prefrontal cortex in cognitive control and executive function. Neuropsychopharmacology 47, 72–89 (2022).

92. Snyder, H. R., Kaiser, R. H., Warren, S. L. & Heller, W. Obsessive-compulsive disorder is associated with broad impairments in executive function: A meta-analysis. Clin Psychol Sci 3, 301–330 (2015).

93. Vriend, C. et al. Switch the itch: A naturalistic follow-up study on the neural correlates of cognitive flexibility in obsessive-compulsive disorder. Psychiatry Research - Neuroimaging 213, 31–38 (2013).

94. Nambu, A., Tokuno, H. & Takada, M. Functional significance of the cortico-subthalamo-pallidal ‘hyperdirect’ pathway. Neuroscience Research 43, 111–117 (2002).

95. Bari, A. & Robbins, T. W. Inhibition and impulsivity: Behavioral and neural basis of response control. Progress in Neurobiology 108, 44–79 (2013).

96. Rasmussen, J., Siev, J., Abramovitch, A. & Wilhelm, S. Scrupulosity and contamination OCD are not associated with deficits in response inhibition. Journal of Behavior Therapy and Experimental Psychiatry 50, 120–126 (2016).

97. Silveira, V. P. et al. Exploring response inhibition and error monitoring in obsessive-compulsive disorder. J Psychiatr Res. 126, 26–326–33 (2020).

98. Kalanthroff, E. et al. The role of response inhibition in medicated and unmedicated obsessive-compulsive disorder patients: Evidence from the stop-signal task. Depression and Anxiety 34, 301–306 (2017).

99. Rappel, P. et al. Subthalamic theta activity: A novel human subcortical biomarker for obsessive compulsive disorder. Translational Psychiatry 8, (2018).

100. Sildatke, E. et al. Deep Brain Stimulation Reduces Conflict-Related Theta and Error-Related Negativity in Patients With Obsessive–Compulsive Disorder. Neuromodulation: Technology at the Neural Interface 25, 245–252 (2022).

101. Schüller, T. et al. Performance monitoring in obsessive–compulsive disorder: Insights from internal capsule/nucleus accumbens deep brain stimulation. NeuroImage: Clinical 31, 102746 (2021).

102. Murphy, D. L., Timpano, K. R., Wheaton, M. G., Greenberg, B. D. & Miguel, E. C. Obsessive-compulsive disorder and its related disorders: a reappraisal of obsessive-compulsive spectrum concepts. Dialogues in Clinical Neuroscience 12, 131–148 (2010).

103. Bartz, J. A. & Hollander, E. Is obsessive-compulsive disorder an anxiety disorder? Progress in Neuro-Psychopharmacology and Biological Psychiatry 30, 338–352 (2006).

104. Myers-Schulz, B. & Koenigs, M. Functional anatomy of ventromedial prefrontal cortex: Implications for mood and anxiety disorders. Molecular Psychiatry 17, 132–141 (2012).

105. Apergis-Schoute, A. M. et al. Neural basis of impaired safety signaling in Obsessive Compulsive Disorder. Proceedings of the National Academy of Sciences of the United States of America 114, 3216–3221 (2017).

106. Milad, M. R. et al. Deficits in conditioned fear extinction in obsessive-compulsive disorder and neurobiological changes in the fear circuit. JAMA Psychiatry 70, 608–618 (2013).

107. Figee, M. et al. Deep brain stimulation restores frontostriatal network activity in obsessive-compulsive disorder. Nature Neuroscience 16, 386–387 (2013).

108. Yap, K., Mogan, C. & Kyrios, M. Obsessive-compulsive disorder and comorbid depression: The role of OCD-related and non-specific factors. Journal of Anxiety Disorders 26, 565–573 (2012).

109. Drevets, W. C. Orbitofrontal cortex function and structure in depression. Annals of the New York Academy of Sciences 1121, 499–527 (2007).

110. Milad, M. R. & Rauch, S. L. The role of the orbitofrontal cortex in anxiety disorders. Annals of the New York Academy of Sciences 1121, 546–561 (2007).

111. Apergis-Schoute, A. M. et al. Hyperconnectivity of the ventromedial prefrontal cortex in obsessive-compulsive disorder. Brain and Neuroscience Advances 2, 239821281880871 (2018).

112. Figee, M. et al. Dysfunctional reward circuitry in obsessive-compulsive disorder. Biological Psychiatry 69, 867–874 (2011).

113. Kaufmann, C. et al. Medial prefrontal brain activation to anticipated reward and loss in obsessive-compulsive disorder. NeuroImage: Clinical 2, 212–220 (2013).

114. Abramovitch, A., Pizzagalli, D. A., Reuman, L. & Wilhelm, S. Anhedonia in obsessive-compulsive disorder: Beyond comorbid depression. Psychiatry Research 216, 223–229 (2014).

115. Hollunder, B. & Horn, A. Mapping the dysfunctome provides an avenue for targeted brain circuit therapy. Nature Neuroscience 27, 401–402 (2024).

116. Allawala, A. et al. A novel framework for network-targeted neuropsychiatric deep brain stimulation. Neurosurgery 89, 116–121 (2021).

117. Figee, M. & Mayberg, H. The future of personalized brain stimulation. Nature Medicine 27, 196–197 (2021).

118. Davidson, B. et al. Lesional psychiatric neurosurgery: meta-analysis of clinical outcomes using a transdiagnostic approach. J Neurol Neurosurg Psychiatry 93, 207–215 (2022).

119. Yücel, M. et al. A transdiagnostic dimensional approach towards a neuropsychological assessment for addiction: An international Delphi consensus study. Addiction 114, 1095–1109 (2019).

120. Hollunder, B., Ganos, C. & Horn, A. Deep brain stimulation: From sweet spots to sweet networks? Biological Psychiatry: Cognitive Neuroscience and Neuroimaging 6, 939–941 (2021).

121. Butenko, K. et al. Engaging dystonia networks with subthalamic stimulation. Proceedings of the National Academy of Sciences 122, e2417617122 (2025).

122. Ashkan, K., Rogers, P., Bergman, H. & Ughratdar, I. Insights into the mechanisms of deep brain stimulation. Nature Reviews Neurology 13, 548–554 (2017).

123. Neudorfer, C. et al. Kilohertz-frequency stimulation of the nervous system: A review of underlying mechanisms. Brain Stimulation 14, 513–530 (2021).

124. Noecker, A. M. et al. StimVision v2: Examples and applications in subthalamic deep brain stimulation for Parkinson’s disease. Neuromodulation 24, 248–258 (2021).

125. Horn, A. et al. Lead-DBS v2: Towards a comprehensive pipeline for deep brain stimulation imaging. NeuroImage 184, 293–316 (2019).

126. Horn, A. & Fox, M. D. Opportunities of connectomic neuromodulation. NeuroImage 221, 117180 (2020).

127. Faul, F., Erdfelder, E., Lang, A.-G. & Buchner, A. G*Power 3: A flexible statistical power analysis program for the social, behavioral, and biomedical sciences. Behavior Research Methods 39, 3759–175–191 (2007).

128. Faul, F., Erdfelder, E., Buchner, A. & Lang, A.-G. Statistical power analyses using G * Power 3. 1 : Tests for correlation and regression analyses. Behavior Genetics 41, 1149–1160 (2009).

129. Beck, A. T., Ward, C. H., Mendelson, M., Mock, J. & Erbaugh, J. An Inventory for Measuring Depression. 561–571 (1960).

130. Montgomery, S. A. & Asberg, M. A new depression scale designed to be sensitive to change. British Journal of Psychiatry 134, 382–389 (1979).

131. Hamilton, M. A rating scale for depression. J Neurol Neurosurg Psychiatry 23, 56–62 (1960).

132. Hamilton, M. the Assessment of Anxiety States By Rating. British Journal of Medical Psychology 32, 50–55 (1959).

133. Beck, Brown, Epstein, & Steer. An inventory for measuring clinical anxiety: Psychometric properties. Clinical Psychology 56, 893–897 (1988).

134. Spielberger, C. D., Gorsuch, R. L. & Lushene, R. E. Manual for the State-Trait Inventory STAI. (Consulting Psychologist Press, Palo Alto, CA, 1970).

135. Endicott, J., Spitzer, R. L., Fleiss, J. L. & Cohen, J. The global assessment scale. A procedure for measuring overall severity of psychiatric disturbance. Archives of General Psychiatryeneral psychiatry 33, 766–771 (1976).

136. Aas, I. M. Guidelines for rating Global Assessment of Functioning (GAF). Ann Gen Psychiatry 10, 2 (2011).

137. American Psychiatric Association. Diagnostic and Statistical Manual of Mental Disorders (5th *Ed.)*. (Arlington, VA).

138. Siddiqi, S. H. et al. Brain stimulation and brain lesions converge on common causal circuits in neuropsychiatric disease. Nature Human Behaviour 5, 1707–1716 (2021).

139. Meyer, G. M. et al. Deep Brain Stimulation for Obsessive-Compulsive Disorder: Optimal stimulation sites. Biological Psychiatry 10.1016/j.biopsych.2023.12.010 (2023) doi:10.1016/j.biopsych.2023.12.010.

140. Reich, M. M. et al. A brain network for deep brain stimulation induced cognitive decline in Parkinson’s disease. Brain 10.1093/brain/awac012 (2022) doi:10.1093/brain/awac012.

141. Stroop, J. R. Studies of interference in serial verbal reactions. Journal of Experimental Psychology 18, 643–662 (1935).

142. Sahakian, B. J. et al. A comparative study of visuospatial memory and learning in Alzheimer-type dementia and Parkinson’s disease. Brain 111, 695–718 (1988).

143. Arnett, J. A. & Labovitz, S. S. Effect of Physical Layout in Performance of the Trail Making Test. Psychological Assessment 7, 220–221 (1995).

144. Horn, A. & Kühn, A. A. Lead-DBS: A toolbox for deep brain stimulation electrode localizations and visualizations. NeuroImage 107, 127–135 (2015).

145. Neudorfer, C. et al. Lead-DBS v3.0: Mapping deep brain stimulation effects to local anatomy and global networks. NeuroImage 268, 119862 (2023).

146. Avants, B. B., Epstein, C. L., Grossman, M. & Gee, J. C. Symmetric diffeomorphic image registration with cross- correlation: Evaluating automated labeling of elderly and neurodegenerative brain. Medical Image Analysis 12, 26–41 (2008).

147. Husch, A., V. Petersen, M., Gemmar, P., Goncalves, J. & Hertel, F. PaCER - A fully automated method for electrode trajectory and contact reconstruction in deep brain stimulation. NeuroImage: Clinical 17, 80–89 (2018).

148. Vorwerk, J., Oostenveld, R., Piastra, M. C., Magyari, L. & Wolters, C. H. The FieldTrip-SimBio pipeline for EEG forward solutions. BioMedical Engineering Online 17, 37 (2018).

149. Treu, S. et al. Deep brain stimulation: Imaging on a group level. NeuroImage 219, 117018 (2020).

150. Irmen, F. et al. Left prefrontal connectivity links subthalamic stimulation with depressive symptoms. Annals of Neurology 87, 962–975 (2020).

151. Horn, A. et al. Optimal deep brain stimulation sites and networks for cervical vs. generalized dystonia. Proceedings of the National Academy of Sciences 119, e2114985119 (2022).

152. Ríos, A. S. et al. Optimal deep brain stimulation sites and networks for stimulation of the fornix in Alzheimer’s disease. Nature Communications 13, (2022).

