## Supplementary Materials for "A Multi-Symptom Circuit Architecture of Obsessive–Compulsive Disorder"

*(Hollunder et al.)*

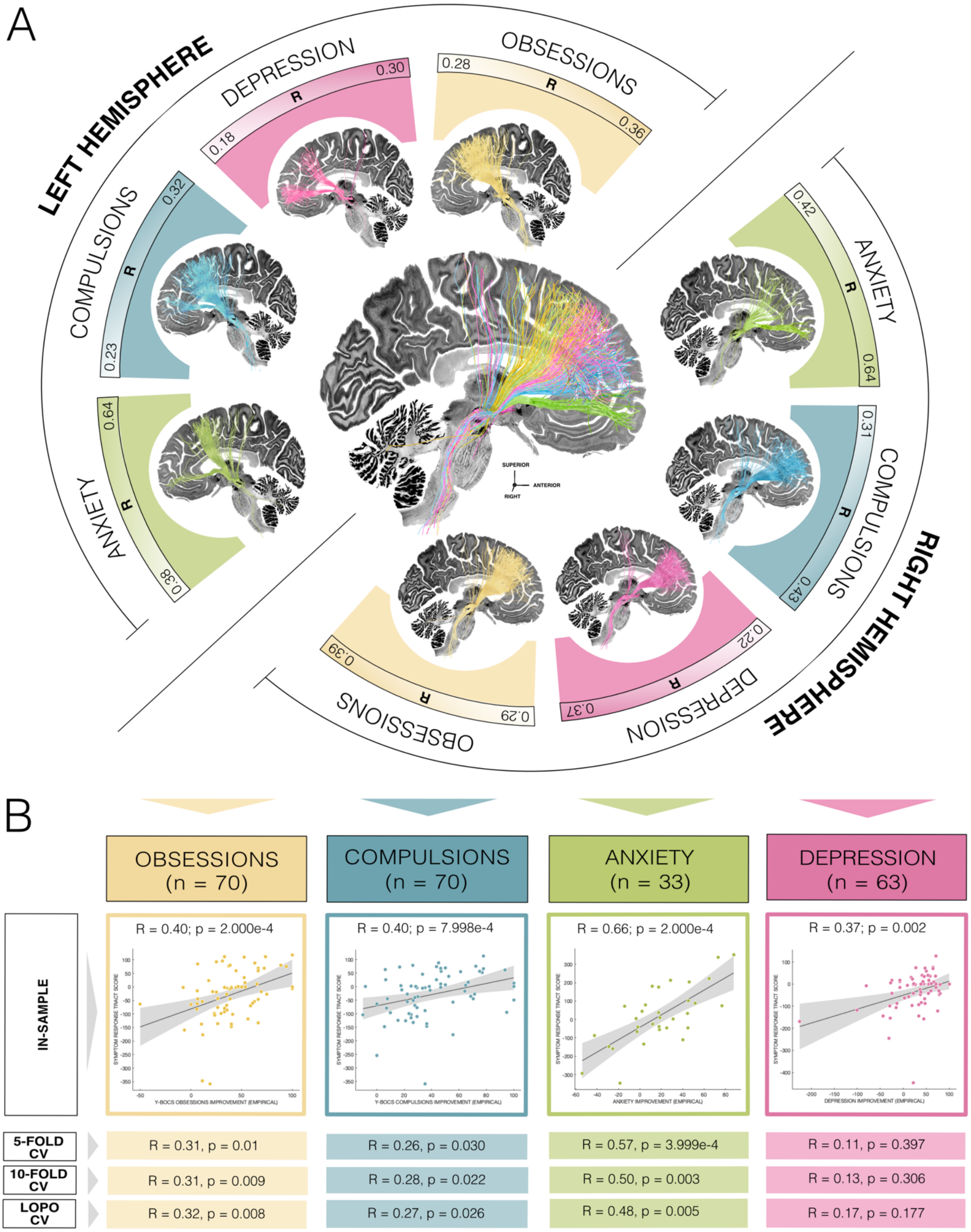


**Fig. S1. Full-sample replication of therapeutic symptom response tracts. (A)** The symptom-associated therapeutic tracts linked to deep brain stimulation (DBS) related improvements in obsessions, compulsions, anxiety, and depression closely mirror those reported in the main manuscript, which were derived from a subset of patients (n = 32) with complete scores across all four symptom domains. Here, tract mappings were generated using the full obsessive-compulsive disorder (OCD) patient cohort (N = 77). However, not all patients had data available for every symptom measure, resulting in variable statistical power depending on data completeness. Again, DBS Fiber Filtering was performed using the normative Human Connectome Project[9] (N = 985) connectome[10] and results were visualized against a sagittal slice (x = -5 mm) of the 100 µm BigBrain template[11]. Tracts are thresholded and color-coded per symptom domain, with intensity scaled by R-values reflecting the importance of tract modulation for symptom improvement. **(B)** Analogous to the main analysis, model validity was assessed by evaluating each symptom tract for its ability to explain outcome variance within the cohort (in-sample correlation). The plot shows the relationship of the average bilateral Symptom Response Tract Score (reflecting the extent of tract modulation across bihemispheric stimulation fields per patient) with observed symptom changes across the patient sample. To ensure generalizability, several cross-validation (CV) strategies were applied, including 5-fold, 10-fold, and leave-one-patient-out (LOPO) designs, using subsets of the data to predict outcomes in held-out cases. Gray shaded regions indicate 95% confidence intervals.

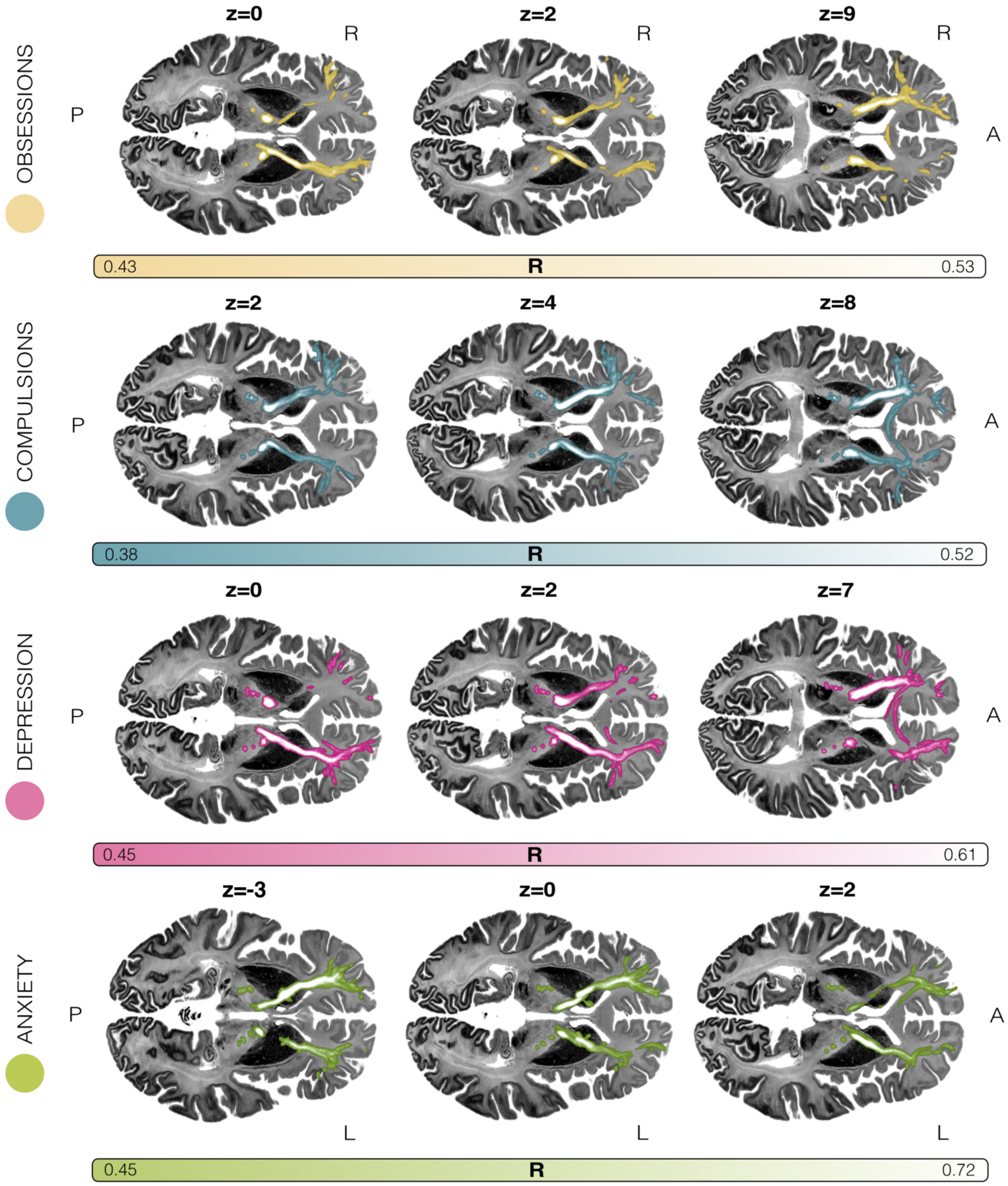


**Fig. S2: Axial 2D representations of core symptom response tracts.** For increased insight into their anatomical course, 2D views of deep brain stimulation (DBS) related symptom response tracts for obsessions, compulsions, depression and anxiety symptoms are displayed in symptom-specific color against different axial slices of the BigBrain template[11] in ICBM 2009b Nonlinear Asymmetric ('MNI') space and in radiological convention. Note that, critically, the z-slice heights vary across rows, chosen to best visualize each tract. For accurate interpretation, the tract views represented here should be compared with the main figures. These tracts can be seen as models of how DBS electrodes should be ideally connected to maximize symptom relief in each respective domain. They were derived by means of DBS Fiber Filtering, integrating symptom-specific imaging, stimulation information and clinical outcome scores with a normative group connectome calculated from 985 participants of the Human Connectome Project[9].

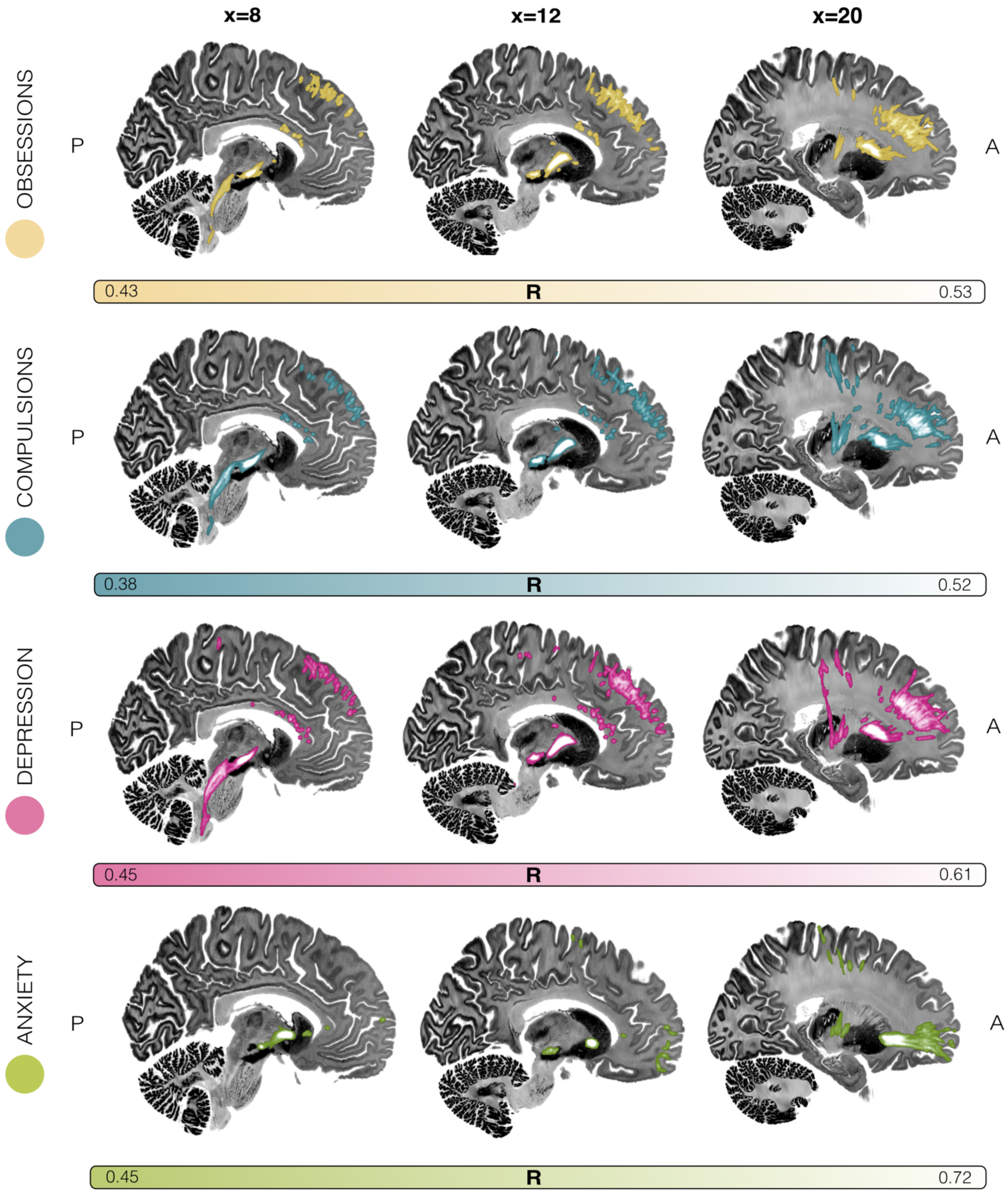


**Fig. S3: Sagittal 2D representations of core symptom response tracts.** The trajectories of neural tracts associated with clinical improvement in obsessions, compulsions, depression, and anxiety following deep brain stimulation (DBS), each rendered in a distinct color and in different 2D sagittal views. These fiber pathways are visualized against slices of the BigBrain template[11] and mapped in standard template space. These tract models represent optimal connections between DBS electrodes and brain regions relevant for alleviation across the respective obsessive-compulsive disorder (OCD) symptom domains. Their identification involved a technique termed DBS Fiber Filtering and leveraged a normative structural connectome constructed from diffusion-weighted imaging data of 985 individuals from the Human Connectome Project[9], enabling a probabilistic estimation of the most therapeutically relevant white matter circuits.

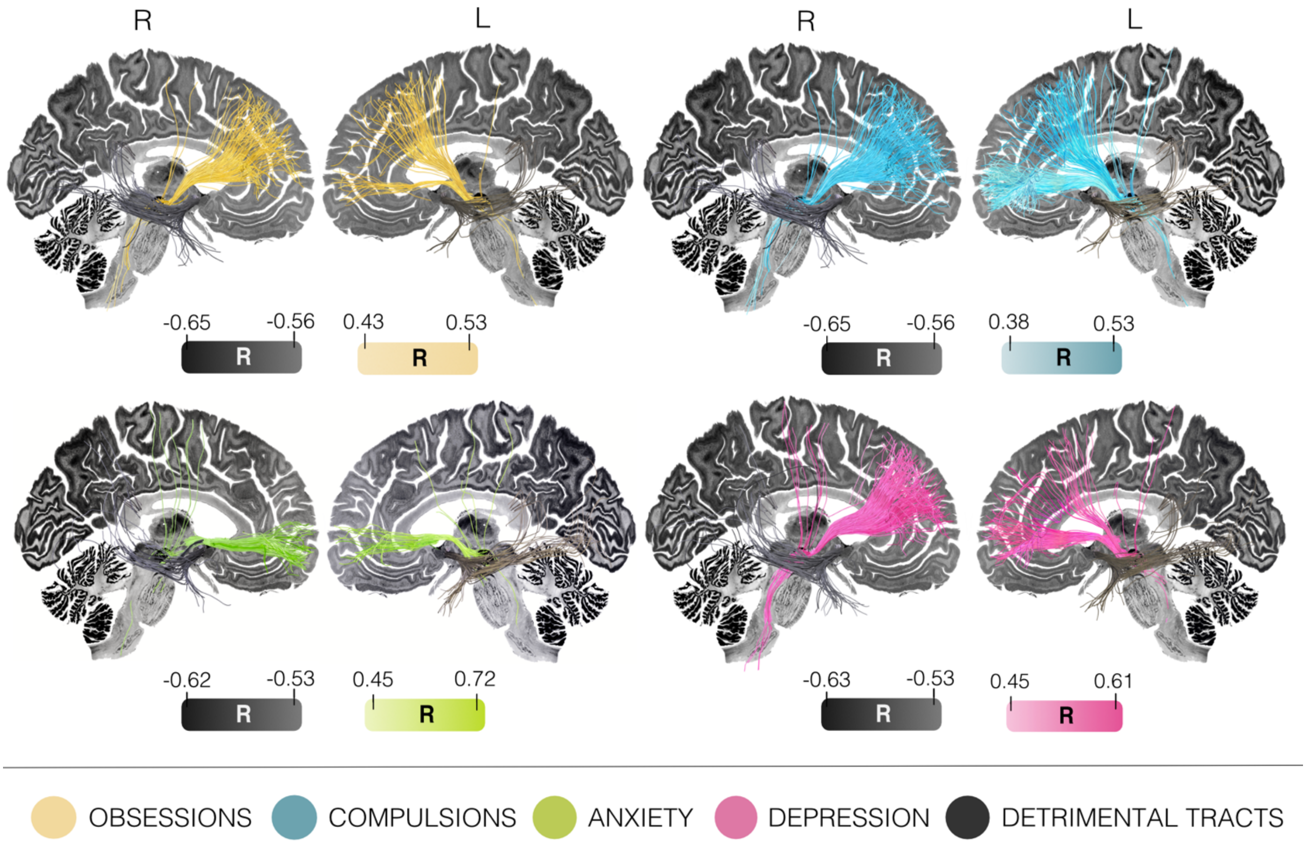


**Fig. S4: Optimal and detrimental symptom response tracts in deep brain stimulation (DBS) for obsessive-compulsive disorder (OCD).** Electrode connectivity associated with maximized improvement (in symptom-specific color) and worsening (in black) following DBS in obsessions (yellow), compulsions (blue), anxiety (green), and depression (pink), based on the subset of patients with OCD featuring a complete set of assessments in all four core symptoms (n = 32). Each tract in these symptom profiles was tagged and colored by an R-value, coding for the Spearman's correlation of its stimulation intensity with clinical improvements in the respective symptom across the patient cohort. Results are informed on a population-based group connectome[10] of 985 healthy adults from the Human Connectome Project (HCP)[9]. Symptom-wise results are represented against the backdrop of a sagittal slice (x = -5 mm) extracted from the Big Brain template[11].

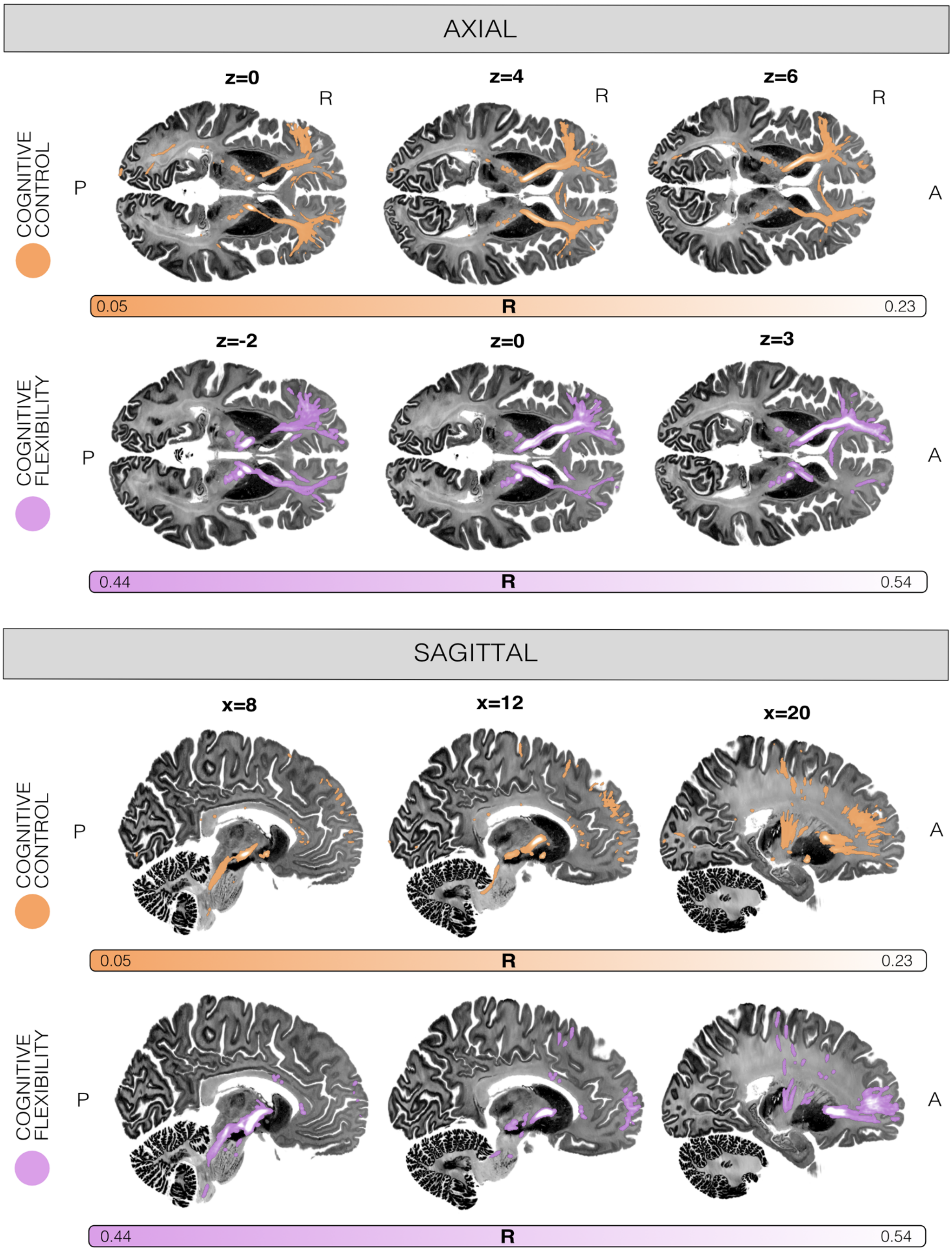


**Fig. S5. Axial and sagittal representations of white matter pathways associated with improved cognitive control and cognitive flexibility deficits.** Neural tracts linked to clinical improvement in symptoms related to impaired cognitive control and cognitive flexibility in obsessive-compulsive disorder following deep brain stimulation (DBS) are shown in distinct colors across axial and sagittal views. Note that the z-slice heights are intentionally not uniform across rows, but each is adjusted to best showcase the respective tract. Interpretation of tracts should take main figures into consideration. These pathways are visualized using the high-resolution BigBrain template[11]. The tract models illustrate optimal connections between DBS electrode sites and key brain regions implicated in these cognitive domains. Their identification was achieved through DBS Fiber Filtering, using a normative structural connectome derived from diffusion-weighted imaging data of 985 participants in the Human Connectome Project.

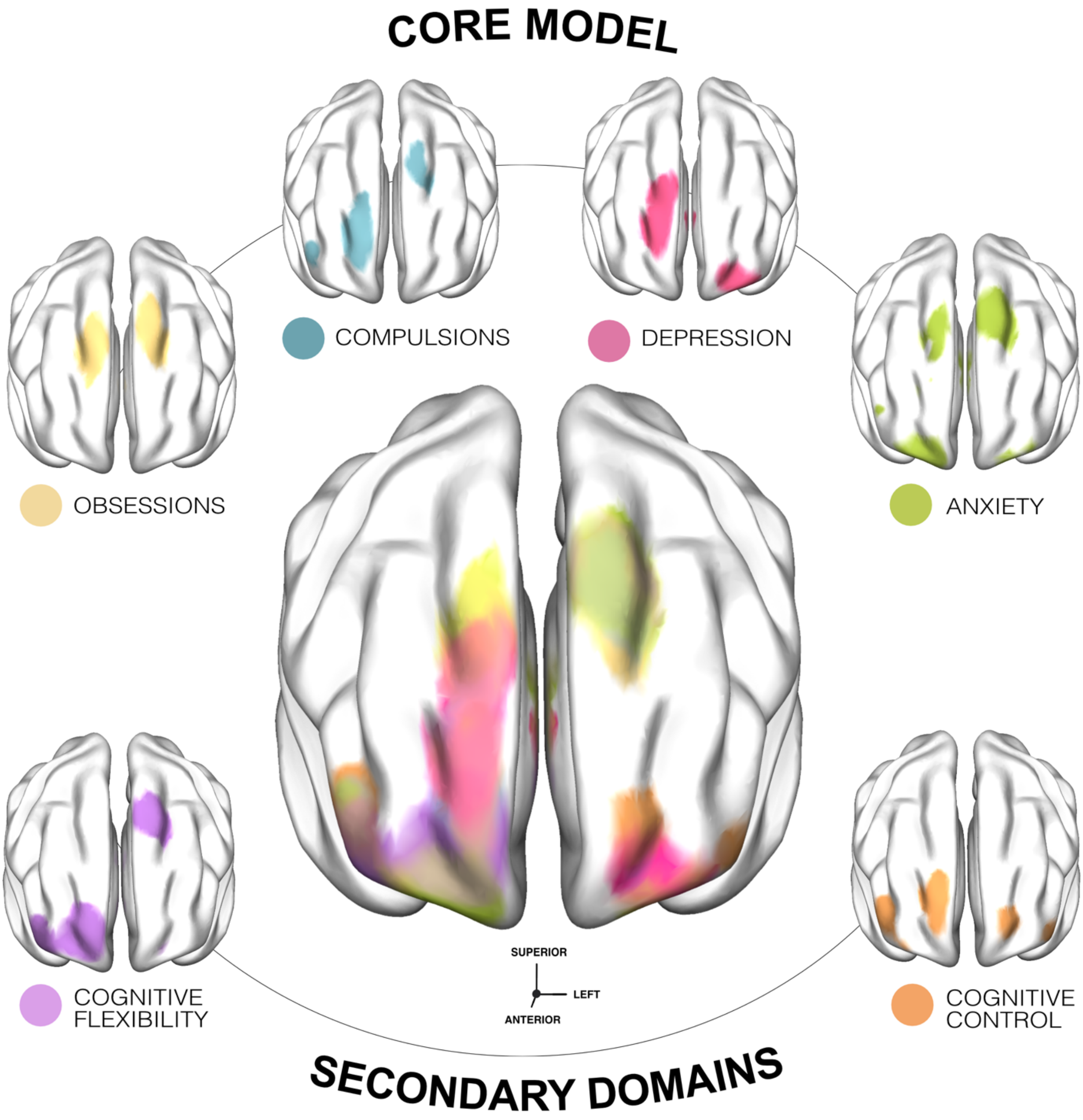


**Fig. S6. Cortical topography of different symptom response tracts in obsessive-compulsive disorder (OCD).** To determine the cortical topography of optimal symptom response tract mappings in the full OCD cohort (N = 77), density maps are overlayed on top of a brain template in ICBM 2009b Nonlinear Asymmetric ('MNI')

space. The mappings are displayed in a smoothed, thresholded, and binarized format, with colors representing symptom-specific hues. The upper row displays core symptom models, whereas the bottom row shows more explorative symptom domains with weaker evidence due to a lower n.

**Table S1**. Demographic and clinical characteristics of obsessive-compulsive disorder cohorts.

| Cohort | Cologne | Boston | Brisbane | London | Madrid | Melbourne | Toronto | Grenoble | Full sample |
|---|---|---|---|---|---|---|---|---|---|
| Surgical center | University Hospital Cologne | Massachusetts General Hospital. Boston | St Andrew's War Memorial Hospital, Brisbane | National Hospital for Neurology and Neurosurgery, London | Hospital Clínico San Carlos, Madrid | St. Vincent's Hospital, Melbourne | Toronto Western Hospital, Toronto | University Hospital Grenoble | - |
| DBS target | ALIC | ALIC | BNST | VC/VS & STN | NAcc | NAcc | ITP | STN | ALIC (n = 29), NAcc (n = 15), BNST (n = 9), ITP (n = 5), STN (n = 13), VC/VS & STN (n = 6) |
| Average MNI coordinate of active contact (x,y,z) | L: -10.8, 6.5, -6.3;<br>R: 10.7, 6.4, -5.5 | L: -13.7, 5.1, -0.5;<br>R: 14.1, 4.2, -2.4 | L: -7.3, 1.6, -9.5;<br>R: 7.0, 1.2, -8.9 | ALIC – L: -14.9, 9.2, -2.4;<br>R: 12.5, 9.9, -2.5;<br>STN – L: -9.8, -12.8, -9.1;<br>R: 8.9, -12.2, -8.2 | L: -11.5, 11.9, 1.8;<br>R: 10.5, 11.6, 0.4 | L: -11.3, 8.8, -5.4;<br>R:10.8, 9.1, -6.8 | L: -6.4, -0.5, -3.5;<br>R: 5.7, -0.6, -4.7 | L: -10.1, -12.0, -7.8;<br>R: 10.4, -11.1, -7.4 | ALIC – L: -12.1, 6.7, -4.5; R: 11.7, 6.5, -4.3;<br>NAcc – L: -11.4, 10.3, -2.0; R: 10.6, 10.2, -3.5;<br>BNST – L: -7.3, 1.6, -9.5; R: 7.0, 1.2, -8.9;<br>ITP – L: -6.4, -0.5, -3.5; R: 5.7, -0.6, -4.7;<br>STN – L: -10.0, -12.2, -8.2; R: 9.9, -11.4, -7.6 |
| Number of patients | n = 22 | n = 7 | n = 9 | n = 6 | n = 7 | n = 8 | n = 5 | n = 13 | N = 77 |
| Time at FU (in months) | 12 | 18 | 12 | 6 | median = 15 | 12 | 12 | 24 | median = 12 |
| Related citation | Baldermann *et al.*[1] | McLaughlin *et al.*[2] | Mosley *et al.*[3] | Tyagi *et al.*[4] | Barcia *et al.*[5] | Acevedo *et al.*[6] | Lee *et al.*[7] | Polosan *et al.*[8] | - |
| **Imaging and electrode specifications** | | | | | | | | | |
| Postoperative imaging modality | CT (n = 22) | CT (n = 7) | CT (n = 9) | MRI (n = 6) | CT (n = 7) | CT (n = 8) | MRI (n = 5) | MRI (n = 10), CT (n = 3) | MRI (n = 21), CT (n = 56) |
| Electrode models | MDT 3387 (n = 3) or MDT 3389 (n = 19) | MDT 3387 (n = 7) | MDT 3389 (n = 9) | STN: MDT 3389 (n = 6); VC/VS: MDT 3387 (n = 6) | MDT 3391 (n = 7) | MDT 3389 (n = 8) | St. Jude 6143 ANS (n = 3), MDT 3387 (n = 2) | MDT 3389 (n = 13) | MDT 3387 (n = 18), MDT 3389 (n=55), MDT 3391 (n = 7), St. Jude 6143 ANS (n = 3) |
| **Demographics** | | | | | | | | | |
| Sex (M/F) | 9M/13F | 3M/4F | 5M/4F | 5M/1F | 4M/3F | 5M/3F | 2M/3F | 4M/9F | 37M/40F |
| Age at onset (mean ± SD, in years) | 16.9 ± 11.6 | 13.7 ± 4.5 | 9.7 ± 4.7 | 21.3 ± 8.3 | 10.7 ± 5.1 | 14.2 ± 6.9 | 16.2 ± 6.7 | 20.2 ± 7.8 | 17.8 ± 10.2 |
| Age at surgery (mean ± SD, in years) | 41.7 ± 12.6 | 38.9 ± 16.8 | 47.7 ± 11.4 | 45.5 ± 10.5 | 33.3 ± 9.5 | 37.8 ± 9.2 | 32.4 ± 9.5 | 39.2 ± 8.2 | 38.2 ± 14.2 |
| **Clinical symptom assessments** | | | | | | | | | |
| **Y-BOCS – Total** | | | | | | | | | |
| Observations | n = 22 | n = 7 | n = 9 | n = 6 | n = 7 | n = 8 | n = 5 | n = 13 | N = 77 |
| Baseline (mean ± SD) | 31.4 ± 4.2 | 33.6 ± 2.4 | 32.7 ± 2.7 | 36.2 ± 1.8 | 29.4 ± 8.6 | 29.8 ± 3.8 | 35.0 ± 2.3 | 33.5 ± 3.8 | 32.4 ± 4.5 |
| FU (mean ± SD) | 21.8 ± 7.7 | 20.3 ± 10.4 | 19.3 ± 5.9 | 14.3 ± 4.1 | 16.1 ± 6.0 | 18.6 ± 10.1 | 16.8 ± 4.9 | 16.9 ± 9.7 | 18.8 ± 8.0 |

| | | | | | | | | | |
|---|---|---|---|---|---|---|---|---|---|
| Improvement (%) | 31.0 ± 20.5 | 40.1 ± 29.8 | 40.2 ± 19.2 | 60.0 ± 12.6 | 45.9 ± 11.1 | 37.2 ± 34.6 | 52.0 ± 14.1 | 49.6 ± 28.8 | 41.6 ± 24.0 |
| **Y-BOCS – Obsessions** | | | | | | | | | |
| Observations | n = 22 | n = 7 | n = 9 | n = 6 | n = 5 | n = 8 | - | n = 13 | n = 70 |
| Baseline (mean ± SD) | 16.2 ± 2.0 | 16.7 ± 1.4 | 15.8 ± 1.5 | 18.5 ± 0.5 | 13.6 ± 5.5 | 15.1 ± 2.0 | - | 16.4 ± 2.1 | 16.1 ± 2.4 |
| FU (mean ± SD) | 11.0 ± 4.0 | 11.6 ± 5.7 | 9.1 ± 2.0 | 7.2 ± 2.4 | 8.2 ± 1.5 | 9.5 ± 5.4 | - | 7.9 ± 4.9 | 9.5 ± 4.2 |
| Improvement (%) | 32.3 ± 22.4 | 31.6 ± 32.3 | 41.8 ± 13.5 | 61.2 ± 13.3 | 26.2 ± 42.8 | 37.2 ± 35.3 | - | 51.7 ± 30.0 | 39.7 ± 27.8 |
| **Y-BOCS – Compulsions** | | | | | | | | | |
| Observations | n = 22 | n = 7 | n = 9 | n = 6 | n = 5 | n = 8 | - | n = 13 | n = 70 |
| Baseline (mean ± SD) | 15.2 ± 3.2 | 16.9 ± 1.1 | 16.9 ± 2.1 | 17.7 ± 1.9 | 14.7 ± 4.8 | 14.6 ± 2.1 | - | 17.2 ± 2.1 | 12.9 ± 6.0 |
| FU (mean ± SD) | 10.8 ± 3.9 | 8.7 ± 6.3 | 10.2 ± 4.2 | 7.2 ± 1.8 | 6.2 ± 4.5 | 9.1 ± 4.8 | - | 9.0 ± 5.0 | 16.1 ± 17.0 |
| Improvement (%) | 28.4 ± 21.9 | 49.0 ± 36.0 | 38.3 ± 25.2 | 58.6 ± 13.1 | 60.0 ± 25.3 | 37.2 ± 34.5 | - | 47.6 ± 29.0 | 41.2 ± 27.7 |
| **Depression** | | | | | | | | | |
| Assessment (Observations) | BDI (n = 17) | MADRS (n = 7) | MADRS (n = 9) | BDI (n = 6) | BDI (n = 6) | HAM-D (n = 6) | HAM-D (n = 5) | HAM-D (n = 6), MADRS (n = 1) | n = 63 (BDI: n = 29, HAM-D: n = 17, MADRS: n = 17) |
| Baseline (mean ± SD) | 21.8 ± 11.2 | 34.0 ± 5.6 | 21.2 ± 9.5 | 37.7 ± 13.4 | 30.7 ± 9.8 | 17.7 ± 6.3 | 14.0 ± 7.2 | HAM-D: 12.5 ± 3.6, MADRS: 5 | BDI: 26.4 ± 12.8, HAM-D: 14.8 ± 5.9, MADRS: 25.5 ± 11.2 |
| FU (mean ± SD) | 19.4 ± 12.5 | 20.9 ± 11.5 | 10.8 ± 7.7 | 17.0 ± 7.6 | 19.0 ± 10.8 | 12.5 ± 5.2 | 6.4 ± 3.4 | HAM-D: 8.7 ± 3.4, MADRS: 4 | BDI: 18.8 ± 11.0, HAM-D: 9.4 ± 4.6, MADRS: 14.5 ± 10.5 |
| Improvement (%) | -13.5 ± 70.0 | 37.5 ± 35.5 | 54.4 ± 21.6 | 54.5 ± 17.4 | 35.1 ± 36.5 | 30.6 ± 10.4 | 33.6 ± 67.7 | 25.9 ± 31.5 | 25.3 ± 51.3 |
| **Anxiety** | | | | | | | | | |
| Assessment (Observations) | STAI – State section (n = 16) | - | - | BAI (n = 6) | HAM-A (n = 5) | HAM-A (n = 6) | - | - | n = 33 (STAI – State section: n = 16, BAI: n = 6, HAM-A: n = 11) |
| Baseline (mean ± SD) | 53.6 ± 12.1 | - | - | 40.0 ± 8.7 | 22.8 ± 7.3 | 20.7 ± 9.5 | - | - | STAI – State section: 53.6 ± 12.1, BAI: 40.0 ± 8.7, HAM-A: 21.6 ± 8.2 |
| FU (mean ± SD) | 53.4 ± 14.9 | - | - | 18.2 ± 9.4 | 21.2 ± 10.2 | 10.5 ± 4 | - | - | STAI – State section: 53.4 ± 14.9, BAI: 18.2 ± 9.4, HAM-A: 15.4 ± 9.0 |
| Improvement (%) | -1.3 ± 26.6 | - | - | 50.2 ± 30.7 | 9.9 ± 24.6 | 43.3 ± 20.1 | - | - | 17.9 ± 33.6 |
| **Global Assessment of Functioning** | | | | | | | | | |
| Observations | n = 17 | - | - | n = 6 | n = 6 | n = 4 | - | n = 11 | n = 44 |
| Baseline (mean ± SD) | 35.4 ± 2.9 | - | - | 22.0 ± 9.8 | 35.0 ± 16.4 | 43.3 ± 9.3 | - | 34.2 ± 3.8 | 34.0 ± 9.2 |
| FU (mean ± SD) | 55.0 ± 11.9 | - | - | 67.5 ± 11.7 | 49.2 ± 22.2 | 56.5 ± 9.3 | - | 66.3 ± 20.5 | 58.9 ± 16.5 |
| Improvement (%) | 57.3 ± 41.1 | - | - | 365.2 ± 465.6 | 80.8 ± 112.5 | 39.5 ± 58.4 | - | 95.4 ± 62.3 | 110.4 ± 198.2 |
| **Cognitive control** | | | | | | | | | |
| Assessment (Observations) | Stroop (speed, incongruent | - | - | - | Stroop (correct answers / 1 min., | - | - | - | n = 23 |

| | | | | | | | | | |
|---|---|---|---|---|---|---|---|---|---|
| | condition; n = 16) | | | | incongruent condition; n = 7) | | | | |
| Baseline (mean ± SD) | 101.2 ± 37.1 sec. | - | - | - | 31.1 ± 9.5 | - | - | - | Stroop (correct answers / 1 min., incongruent condition): 31.1 ± 9.5 words, Stroop (speed, incongruent condition): 101.2 ± 37.1 sec. |
| FU (mean ± SD) | 91.4 ± 40.5 sec. | - | - | - | 39.7 ± 11.0 | - | - | - | Stroop (correct answers / 1 min., incongruent condition): 39.7 ± 11.0, Stroop (speed, incongruent condition): 91.4 ± 40.5 sec. |
| Improvement (%) | 7.6 ± 25.7 | - | - | - | 35.3 ± 55.5 | - | - | - | 16.0 ± 38.2 |
| **Cognitive flexibility** | | | | | | | | | |
| Assessment (Observations) | - | - | - | EDS (log errors; n = 6) | TMT-B (time; n = 7) | TMT-B (time; n = 5) | - | - | n = 18 |
| Baseline (mean ± SD) | - | - | - | 1.0 ± 0.4 | 161.0 ± 63.6 sec. | 71.2 ± 9.6 sec. | - | - | TMT-B (time): 123.6 ± 66.1 sec., EDS (log errors): 1.0 ± 0.4 |
| FU (mean ± SD) | - | - | - | 0.5 ± 0.5 | 152.6 ± 148.5 sec. | 83.8 ± 36.5 sec. | - | - | TMT-B (time): 123.9 ± 117.4 sec., EDS (log errors): 0.5 ± 0.5 |
| Improvement (%) | - | - | - | 38.8 ± 53.0 | 6.3 ± 73.1 | -18.4 ± 50.1 | - | - | 10.3 ± 61.9 |

*Notes.* Patients in the London cohort received deep brain stimulation (DBS) implants targeting both the ventral capsule/ventral striatum (VC/VS) and the subthalamic nucleus (STN) regions. The clinical trial included separate phases of VC/VS stimulation, STN stimulation, and combined stimulation. This study focuses on clinical outcomes measured during the combined VC/VS and STN stimulation phase. *Abbreviations.* ALIC, Anterior Limb of the Internal Capsule; BAI, Beck Anxiety Inventory; BDI, Beck Depression Inventory; BNST, Bed Nucleus of the Stria Terminalis; CT, computed tomography; EDS, extra-dimensional stage (stage eight) of the Intra-Extra Dimensional Set Shifting subtask of the Cambridge Neuropsychological Test Automated Battery; F, female; FU, follow-up; HAM-A, Hamilton Anxiety Rating Scale; HAM-D, Hamilton Depression Rating Scale; ITP, inferior thalamic peduncle; MADRS, Montgomery-Åsberg Depression Rating Scale; M, male; MDT 3387/3389/3391, Medtronic 3387/3389/3391; MRI, magnetic resonance imaging; NAcc, Nucleus Accumbens; SD, standard deviation; STAI, State-Trait Anxiety Inventory; TMT-B, Trail Making Test – Part B; Y-BOCS, Yale-Brown Obsessive-Compulsive Scale.

**Table S2**. Demographic and clinical characteristics of Tourette syndrome validation cohorts.

| Cohort | Milan | Milan | Maastricht | London | New York | San Francisco | Gainesville | Full sample |
|---|---|---|---|---|---|---|---|---|
| Surgical center | IRCCS Istituto Ortopedico Galeazzi, Milan | IRCCS Istituto Ortopedico Galeazzi, Milan | Maastricht University Medical Center | National Hospital for Neurology and Neurosurgery, London | New York University | University of California San Francisco | University of Florida, Gainesville | - |
| DBS target | amGpi | cmThal | amGpi | amGpi | cmThal | cmThal | cmThal | amGPi (n = 24), cmThal (n = 15) |
| Average MNI coordinate of active contact (x,y,z) | L: -13.6, -0.1, -1.9; R: 14.1, 0.0, -3.0 | L: -7.0, -12.5, -0.1; R: 6.6, -11.8, 0.4 | L: -15.5, -0.9, -1.5; R: 17.0, -2.6, 0.2 | L:-16.4, -1.8, -3.8; R: 16.0, -1.0, -3.0 | L: -9.0, -12.4, -0.6; R: 7.6, -13.5, -0.4 | L: -6.9, -11.7, 0.8; R: 5.4, -11.8, 2.2 | L: -8.1, -13.5, 1.3; R: 7.0, -13.1, 0.6 | amGPi – L: -15.4, -1.2, -3.0; R: 15.5, -0.8, -2.8; cmThal – L: -8.0, -12.7, 0.2; R: 6.9, -12.7, 0.4 |
| Number of patients | n = 9 | n = 6 | n = 2 | n = 13 | n = 3 | n = 1 | n = 5 | N = 39 |
| Time at FU (median, in months) | 6 | 12 | 31.5 | 15 | 10 | 12 | 12 | 12 |
| **Imaging and electrode specifications** | | | | | | | | |
| Postoperative imaging modality | MRI (n = 7), CT (n = 2) | MRI (n = 6) | CT (n = 2) | MRI (n = 13) | CT (n = 3) | MRI (n = 1) | CT (n = 5) | MRI (n = 27), CT (n = 12) |
| Electrode models | MDT 3387 (n = 4), MDT 3389 (n = 5) | MDT 3387 (n = 6) | MDT 3387 (n = 1), MDT 3389 (n = 1) | MDT 3387 (n = 1), MDT 3389 (n = 12) | MDT 3387 (n = 3) | MDT 3387 (n = 1) | NP 3.5 (n = 5) | MDT 3387 (n = 16), MDT 3389 (n = 18), NP 3.5 (n = 5) |
| **Demographics** | | | | | | | | |
| Sex (M/F) | 6M/3F | 5M/1F | 2F | 10M/3F | 2M/1F | 1M | 2M/3F | 26M/13F |
| Age at onset (mean ± SD, in years) | NA | NA | 6 ± 1.4 | 7.2 ± 2.5 | NA | 7 | 7.8 ± 6.3 | 7.2 ± 3.5 |
| Age at surgery (mean ± SD, in years) | 28.2 ± 9.1 | 33.3 ± 6 | 35.0 ± 8.5 | 30.7 ± 10.1 | 19.3 ± 3.1 | 14 | 34.2 ± 3.7 | 29.9 ± 9.0 |
| **Clinical symptom assessments** | | | | | | | | |
| **YGTSS – Total** | | | | | | | | |
| Observations | n = 9 | n = 6 | n = 2 | n = 13 | n = 3 | n = 1 | n = 5 | N = 39 |
| Baseline (mean ± SD) | 74.7 ± 27.8 | 63.0 ± 13.3 | 39.0 ± 4.2 | 59.5 ± 19.2 | 80.3 ± 25.4 | 86 | 91.8 ± 9.6 | 68.9 ± 22.9 |
| FU (mean ± SD) | 36.3 ± 19.1 | 44.5 ± 14.9 | 11.0 ± 2.8 | 41.5 ± 16.6 | 35.3 ± 31.9 | 99 | 65.2 ± 20.7 | 43.3 ± 22.6 |
| Improvement (%) | 32.8 ± 62.8 | 30.4 ± 12.3 | 71.2 ± 10.4 | 30.2 ± 16.9 | 44.0 ± 61.4 | -15.1 | 29.1 ± 21.6 | 32.7 ± 36.7 |
| **Y-BOCS – Total** | | | | | | | | |
| Observations | n = 9 | n = 6 | n = 2 | n = 13 | n = 3 | n = 1 | n = 5 | N = 39 |
| Baseline (mean ± SD) | 31.6 ± 5.8 | 20.3 ± 6.2 | 25.5 ± 7.8 | 19.6 ± 8.5 | 8.0 ± 1.0 | 23 | 19.2 ± 7.4 | 21.9 ± 9.1 |
| FU (mean ± SD) | 20.8 ± 6.6 | 16.5 ± 4.8 | 6.5 ± 9.2 | 14.8 ± 8.4 | 7.0 ± 6.6 | 26 | 19.6 ± 8.8 | 16.3 ± 8.3 |
| Improvement (%) | 32.7 ± 20.8 | 17.8 ± 17.4 | 67.5 ± 46.0 | 13.2 ± 70.2 | 18.5 ± 74.0 | -13 | -4.1 ± 30.2 | 18.7 ± 49.0 |

*Abbreviations:* amGPi, anteromedial internal pallidum; cmThal, centromedian thalamic region; CT, computed tomography; DBS, deep brain stimulation; F, female; FU, Follow-up; M, male; MDT 3387/3389, Medtronic 3387/3389; MRI, magnetic resonance imaging; NA, Not Available; NP 3.5, NeuroPace DL-344-3.5; SD, standard deviation; Y-BOCS, Yale-Brown Obsessive-Compulsive Scale; YGTSS, Yale Global Tic Severity Scale

### References


1. Baldermann, J. C. *et al.* Connectivity profile predictive of effective deep brain stimulation in obsessive-compulsive disorder. *Biological Psychiatry* **85**, 735–743 (2019).
2. McLaughlin, N. C. R. *et al.* Double blind randomized controlled trial of deep brain stimulation for obsessive-compulsive disorder: Clinical trial design. *Contemporary Clinical Trials Communications* **22**, 100785 (2021).
3. Mosley, P. E. *et al.* A randomised, double-blind, sham-controlled trial of deep brain stimulation of the bed nucleus of the stria terminalis for treatment-resistant obsessive-compulsive disorder. *Translational Psychiatry* **11**, 190 (2021).
4. Tyagi, H. *et al.* A randomized trial directly comparing ventral capsule and anteromedial subthalamic nucleus stimulation in obsessive-compulsive disorder: Clinical and imaging evidence for dissociable effects. *Biological Psychiatry* **85**, 726–734 (2019).
5. Barcia, J. A. *et al.* Personalized striatal targets for deep brain stimulation in obsessive-compulsive disorder. *Brain Stimulation* **12**, 724–734 (2019).
6. Acevedo, N. *et al.* Clinical outcomes of deep brain stimulation for obsessive-compulsive disorder: Insight as a predictor of symptom changes. *Psychiatry Clin Neurosci* **78**, 131–141 (2024).
7. Lee, D. J. *et al.* Inferior thalamic peduncle deep brain stimulation for treatment-refractory obsessive-compulsive disorder: A phase 1 pilot trial. *Brain Stimulation* **12**, 344–352 (2019).
8. Polosan, M. *et al.* Affective modulation of the associative-limbic subthalamic nucleus: deep brain stimulation in obsessive–compulsive disorder. *Translational Psychiatry* **9**, (2019).
9. Van Essen, D. C. *et al.* The WU-Minn Human Connectome Project: An overview. *NeuroImage* **80**, 62–79 (2013).
10. Elias, G. J. B. *et al.* A large normative connectome for exploring the tractographic correlates of focal brain interventions. *Scientific Data* **11**, 1–12 (2024).
11. Amunts, K. *et al.* BigBrain: An ultrahigh-resolution 3D human brain model. *Science* **340**, 1472–1475 (2013).
